# Associations between DNA methylation and cognitive function in early-stage hormone receptor-positive breast cancer patients

**DOI:** 10.1101/2024.11.17.24317299

**Authors:** Shuwei Liu, Dongjing Liu, Catherine M. Bender, Kirk I. Erickson, Susan M. Sereika, John R. Shaffer, Daniel E. Weeks, Yvette P. Conley

**Author notes:** **ORCiDs and email addresses**: Shuwei Liu Dongjing Liu Catherine Bender Kirk Erickson Susan Sereika John Shaffer Daniel Weeks Yvette Conley.

## Abstract

**Background:** Approximately one-third of breast cancer (BC) patients show poorer cognitive function (CF) before receiving adjuvant therapy compared with age-matched healthy controls. However, the biological mechanisms driving CF variation in the context of BC remain unclear. In this study, we aimed to identify genes and biological pathways associated with CF in postmenopausal women with early-stage hormone receptor-positive (HR+) BC using DNA methylation (DNAm) data, a dynamic regulator of gene activity.

**Methods:** Epigenome-wide association studies (EWAS) and differentially methylated region analyses were performed for each CF phenotype (seven objective domains and one subjective phenotype) using DNAm data from whole blood samples (n=109) taken at time of enrollment. Post-EWAS functional analyses were performed to enhance the understanding of the CF-related cytosine-phosphate-guanine (CpG) sites.

**Results:** When adjusting for age, verbal IQ scores, and global DNAm signature, cg10331779 near *CTNND2* (p-value= 9.65 × 10^−9^) and cg25906741 in *MLIP* (p-value= 2.01 × 10^−8^) were associated with processing speed and subjective CF, respectively, while regions in/near *SLC6A11*, *PRKG1/CSTF2T*, and *FAM3B* for processing speed, and regions in/near *PI4KB* and *SGCE/PEG10* for mental flexibility were differentially methylated. In addition, beta-estradiol was identified as a common upstream regulator for all the CF phenotypes, suggesting an essential role of estrogen in explaining variation in CF of HR+ BC patients.

**Conclusions:** In our EWAS of 8 CF phenotypes, we found two epigenome-wide significant signals, one at cg10331779 near *CTNND2* with processing speed and the other at cg25906741 in *MLIP* with subjective CF. We also found three differentially methylated regions associated with processing speed and two associated with mental flexibility. These findings need replication in larger cohorts.

## INTRODUCTION

Breast cancer (BC) is the most commonly diagnosed cancer and the second most common cause of death due to cancer for women in the United States^1,2^. One in eight women will be diagnosed with BC in their lifetime with the median age at diagnosis of 62 when most are postmenopausal^1^. With advancements in cancer treatment, the survival rate of BC has increased in recent years. However, some survivors experience long-term negative effects of cancer or treatments^3^, among them is cancer-related cognitive decline (CRCD)^4,5^. CRCD particularly influences memory and decision-making, which leads to poorer quality of life^5^. Approximately 25% to 75% of BC patients develop symptoms of CRCD and 30%-35% of them show poorer cognitive function (CF) before the beginning of adjuvant therapy compared with age-matched healthy controls^4,6,7^.

Objective and subjective CF in BC patients are complex phenotypes that can be affected by many components besides cancer-related factors, such as genetic predisposition, aging, physical inactivity, and social-demographic factors. Although several potential mechanisms contributing to cognitive impairment in BC patients have been proposed, including oxidative stress, chronic inflammation, and white matter damage, to date there has been no consensus. DNA methylation (DNAm), as a dynamic regulator of gene activity that is sensitive to internal and external environmental and behavioral factors, may provide new insights into the biological mechanisms of CF in BC patients.

In this study, we focus on a cohort of postmenopausal women with early-stage hormone receptor-positive (HR+) BC, a common subtype of BC comprising nearly 80% of the total invasive cases^1,8^. In this subtype, most patients receive adjuvant endocrine therapy (ET). Here, we aim to use epigenome-wide DNAm data to evaluate how the CF and the DNAm signature are related in this cohort and find possible genes and biological pathways associated with CF, which may provide insights into the poor CF observed in BC patients.

## MATERIALS AND METHODS

### Participants and Samples

Participants in this study were a subsample of participants in the randomized controlled trial “Exercise Program in Cancer and Cognition” (EPICC) (NCT02793921) (Table S2). The EPICC study protocol and inclusion/exclusion criteria are detailed elsewhere^9^. Postmenopausal women with early-stage HR+ BC were enrolled from clinical sites in Southwestern Pennsylvania^9,10^. Postmenopausal women with stage I-IIIa HR+ BC who were younger than 80 years old, had ≥8 years of education, and had not initiated adjuvant therapy at the time of recruitment were eligible. The criteria were relaxed during the pandemic to include patients within 2 years of completing primary BC treatment (surgery +/− chemotherapy). Some participants had started ET before their pre-randomization blood sample was collected. All participants provided written informed consent.

Blood samples and CF data were collected at pre-randomization and follow-up timepoints to study how ET and exercise intervention impact CF and the role of DNAm (Figure 1, Figure S1). Here we study only the pre-randomization data because our aim was to determine genetic and biological pathways of variation in CRCD and not to determine the impact of the exercise intervention on CRCD (see Bender et al., 2024^10^).

**Figure 1.**
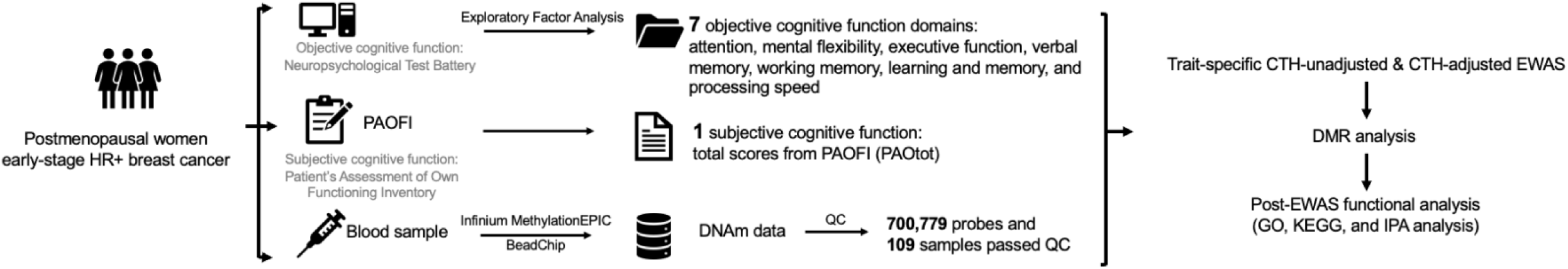
Study workflow

### Phenotypes

#### Objective cognitive function

As previously described^11,12^, a standardized neuropsychological test battery was administered to assess objective CF, yielding multiple measures. These scores were z-transformed with respect to age– and education-matched controls without breast cancer and adjusted so that higher values represent better CF than the controls^10,11^. Seven CF domains were derived using exploratory factor analysis: executive function, attention, mental flexibility, learning and memory, verbal memory, working memory, and processing speed.

#### Subjective cognitive function

Subjective CF was self-assessed using the Patient’s Assessment of Own Functioning Inventory (PAOFI)^13^. The total score based on the sum of the values from 32 items was used in the following analyses, with higher scores indicating worse subjective CF (range 0-160).

#### DNA Methylation Sample Collection

Whole blood samples were collected at pre-randomization and then DNA was extracted using QIAGEN DNA extraction kit. DNA samples were stored in 1X TE buffer at 4℃ for future DNAm data collection. Genome-wide DNAm data were generated by the Infinium MethylationEPIC v1.0 BeadChip (Illumina, San Diego, CA, USA) at the Center for Inherited Disease Research, Johns Hopkins University.

### Quality Control

Quality control of DNAm data was performed by minfi, lumi, Enmix, and ewastools R packages^14–19^. After basic DNAm data processing, DNAm β values and M values were derived as follows: 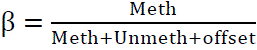 and 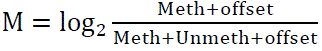 where Meth and Unmeth represent methylated and unmethylated signals, respectively, and the offset term was set as 100. For quality control details, see the Supporting Information. Functional normalization with background and dye bias correction was applied on β values using funNorm^20^. Samples with a detection p-value >0.01 and/or marked as SNP outliers were removed. Technical replicates that had lower detection p-values were retained. After removal of probes with large detection p-values, probes with known SNPs nearby, cross-reactive probes, and Y chromosome probes, 700,779 probes were available (Table S1).

### Statistical Analysis

#### Epigenome-wide association studies (EWAS) and differentially methylated region (DMR) analysis

EWAS were conducted for each phenotype separately using limma R package^21^. As recommended^22^, both EWAS unadjusted for cell type heterogeneity (CTH) (hereinafter referred to as CTH-unadjusted EWAS) and EWAS adjusted for CTH (CTH-adjusted EWAS) were performed. To account for technical artifacts in the CTH-unadjusted analyses, batch, chip row position, median methylated intensities, and median unmethylated intensities were included in the model, in which DNAm M values were regressed on a CF phenotype, as well as age and verbal IQ scores. In CTH-adjusted EWAS, in addition to age and verbal IQ scores, surrogate variables generated by SmartSVA R package were included to control any unwanted variation such as CTH and technical artifacts^22–24^. The EWAS sample sizes varied since only samples with complete observations on CF phenotype and covariates were included (Figure S1). Cytosine-phosphate-guanine (CpG) sites with p-values < 9 × 10^−8^ were regarded as significant differentially methylated CpGs (DMCs) and those with p-values <1 × 10^−5^ were referred to as suggestive DMCs^25^.

DMR analyses were carried out by the dmrff R package using EWAS summary statistics^26^. DMRs that include multiple CpG sites whose DNAm levels were consistently associated with the trait are considered to have better biological meaning than a single CpG site. The dmrff program identified DMRs where the maximum distance between consecutive CpG sites was 500 bp, the EWAS nominal p-value of the CpG sites in the scanned genomic regions was <0.05, and the effect directions of CpG sites in the 500 bp window were consistent. DMRs with a Bonferroni-adjusted p-value <0.05 were considered significant.

#### Post-EWAS functional analysis

To explore which biological processes were associated with CF, we performed gene set enrichment analysis, Gene Ontology (GO) enrichment analysis, and Kyoto Encyclopedia of Genes and Genomes (KEGG) pathway analysis, using the missMethyl R package^27,28^. Limited by the small sample size, we used a less stringent threshold when selecting CpGs as input – CpGs whose nominal p-value was <0.01 in CTH-adjusted EWAS, where the confounding effect of CTH was better controlled, were included in the analysis. GO terms and pathways with a false discovery rate (FDR) <0.05 were considered significant. The same list of CpGs was used to conduct a core analysis in QIAGEN IPA (QIAGEN Inc., https://digitalinsights.qiagen.com/IPA, RRID:SCR_008653) software for pathway analysis and upstream regulators analysis^29^.

*In silico* analyses were then performed on the significant DMCs from CTH-adjusted EWAS. First, a methylation quantitative trait loci (mQTL) lookup analysis was conducted using data from 2,358 blood samples of existing cohorts (TwinsUK and National Child Development Study 1946 and 1958 birth cohorts) based on Infinium MethylationEPIC BeadChip to explore whether the DMCs were under genetic control^30^. Associated SNPs within 1 MB of the selected CpGs were regarded as *cis* mQTLs; others were labeled as *trans* mQTLs. Next, we checked the correlation between DNAm in blood and brain tissues of the selected DMCs. Although blood is the most accessible tissue for generating DNAm data, the brain may be more ideal for studying CF. We queried two databases about the correlation between different brain regions and DNAm in blood generated by Infinium HumanMethylation450 BeadChips and one for DNAm generated by Infinium MethylationEPIC BeadChips^31–33^.

#### Sensitivity analyses

Originally, the EPICC study aimed at recruiting eligible participants who had not started the adjuvant therapy at enrollment^9^. However, due to the COVID-19 pandemic, eligibility criteria were relaxed and some participants had already started ET when their blood samples were collected. Even though ET can improve overall survival for HR+ BC patients, it might be associated with poor CF^34^. To evaluate the relationship between DNAm and CF alone without ET, we performed sensitivity analyses for all the statistical analyses mentioned above on the subset of participants who began ET after blood sample collection.

## RESULTS

### Sample characteristics

Table 1 shows the characteristics of participants included in the analyses. Participants were primarily diagnosed with stage 0 or stage I (77%) BC and self-reported as White (91%) with a mean age of 62 years, a mean estimated verbal IQ score of 112, and a mean educational attainment of 16 years. Compared with education– and age-matched controls without breast cancer, study participants showed poorer CF in many domains, including attention, working memory, verbal memory, and learning and memory. Regarding subjective CF, the mean total scores from self-assessed PAOFI were 20, similar to the average scores for participants without BC in an earlier study^35^. The study participants are similar to the larger EPICC study from which they were recruited (Table S2).

**Table 1.**
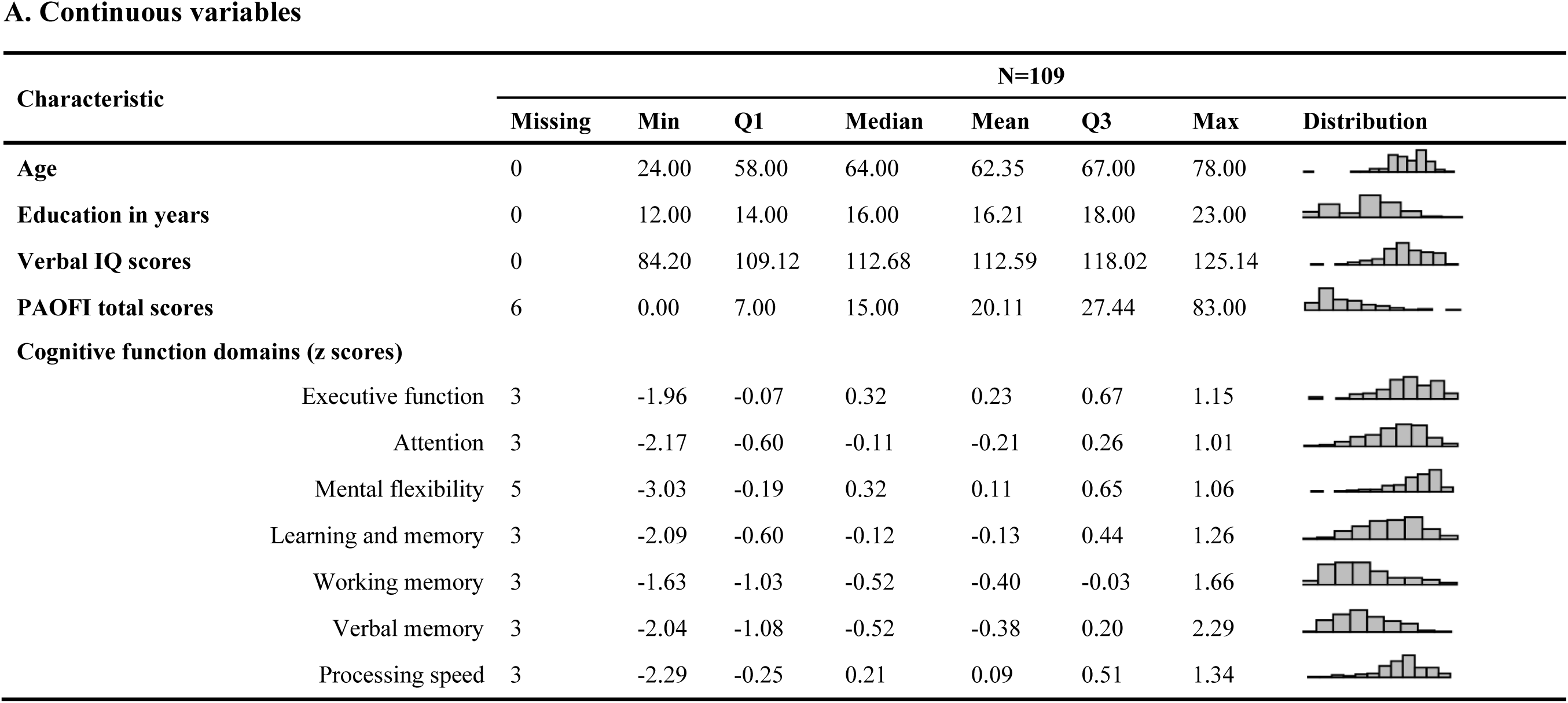
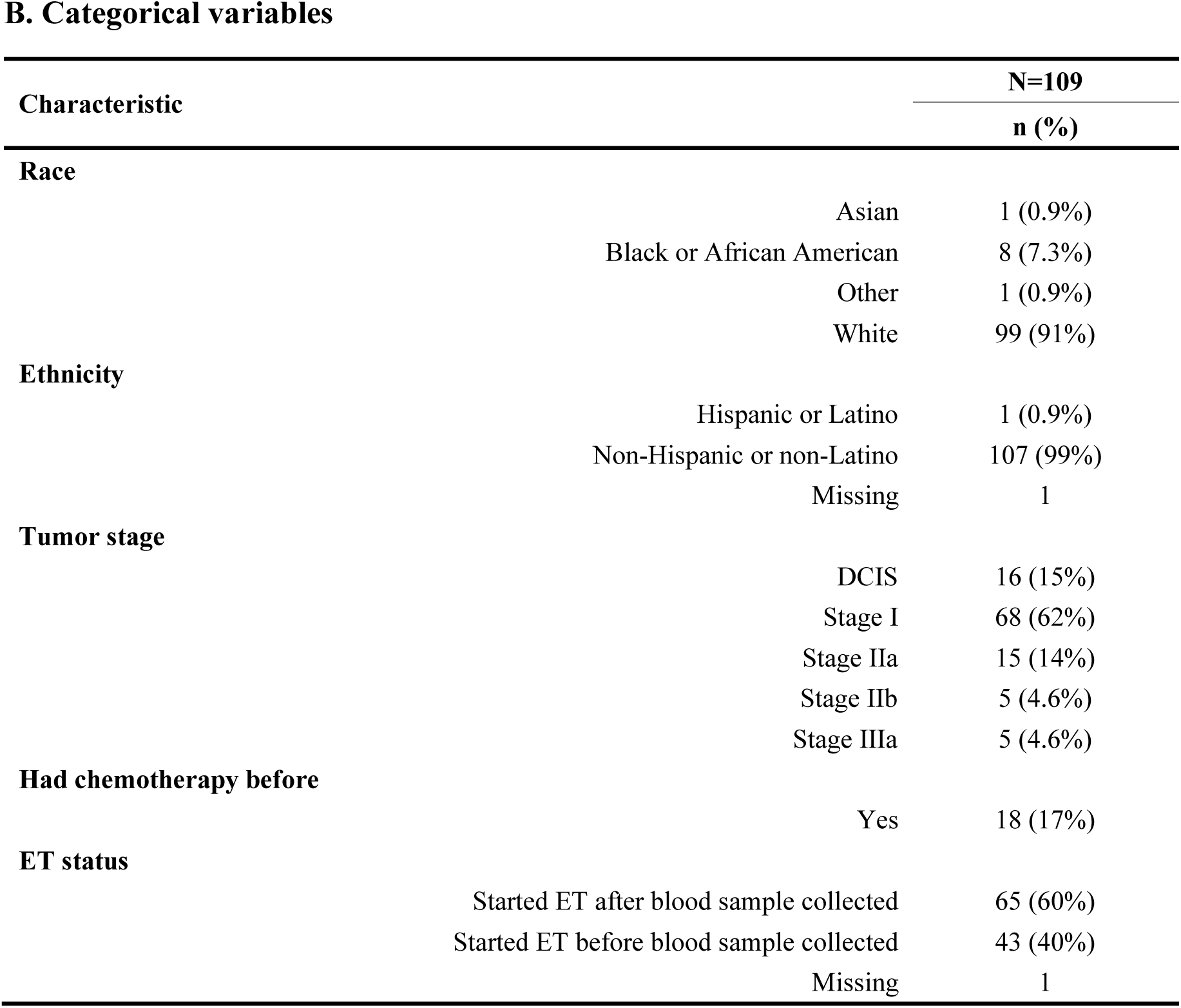
Sample characteristics.

### EWAS and DMR analyses

In CTH-unadjusted analyses, with the genome-wide significance threshold of 9 × 10^−8^, CpG cg08149859 in Pyruvate Dehydrogenase Phosphatase Catalytic Subunit 2 (*PDP2*) was associated with mental flexibility (p-value=4.30 × 10^−8^, coefficient=-0.031), although the association was not significant in the CTH-adjusted EWAS (p-value= 2.27 × 10^−6^, coefficient=-0.024), suggesting that this signal was driven by cell type composition (Table S5). CpG cg24022240 in Adducin 3 (*ADD3*) and cg03444675 near ER Membrane Protein Complex Subunit 2 (*EMC2*) were associated with PAOFI total scores (p-values of 5.23 × 10^−8^ and 8.93 × 10^−8^, respectively). Still, they were not significant after adjusting for CTH (p-values of 8.73 × 10^−7^and 3.40 × 10^−7^, respectively). In the CTH-adjusted analyses, processing speed was significantly associated with cg10331779 in Catenin Delta 2 (*CTNND2*) on chromosome 5 with a p-value of 9.65 × 10^−9^ (Table 2, Figure 2). CpG cg25906741 in Muscular LMNA Interacting Protein (*MLIP*) was negatively associated with PAOFI total scores with a p-value of 2.01 × 10^−8^(Table 2, Figure 3).

**Figure 2.**
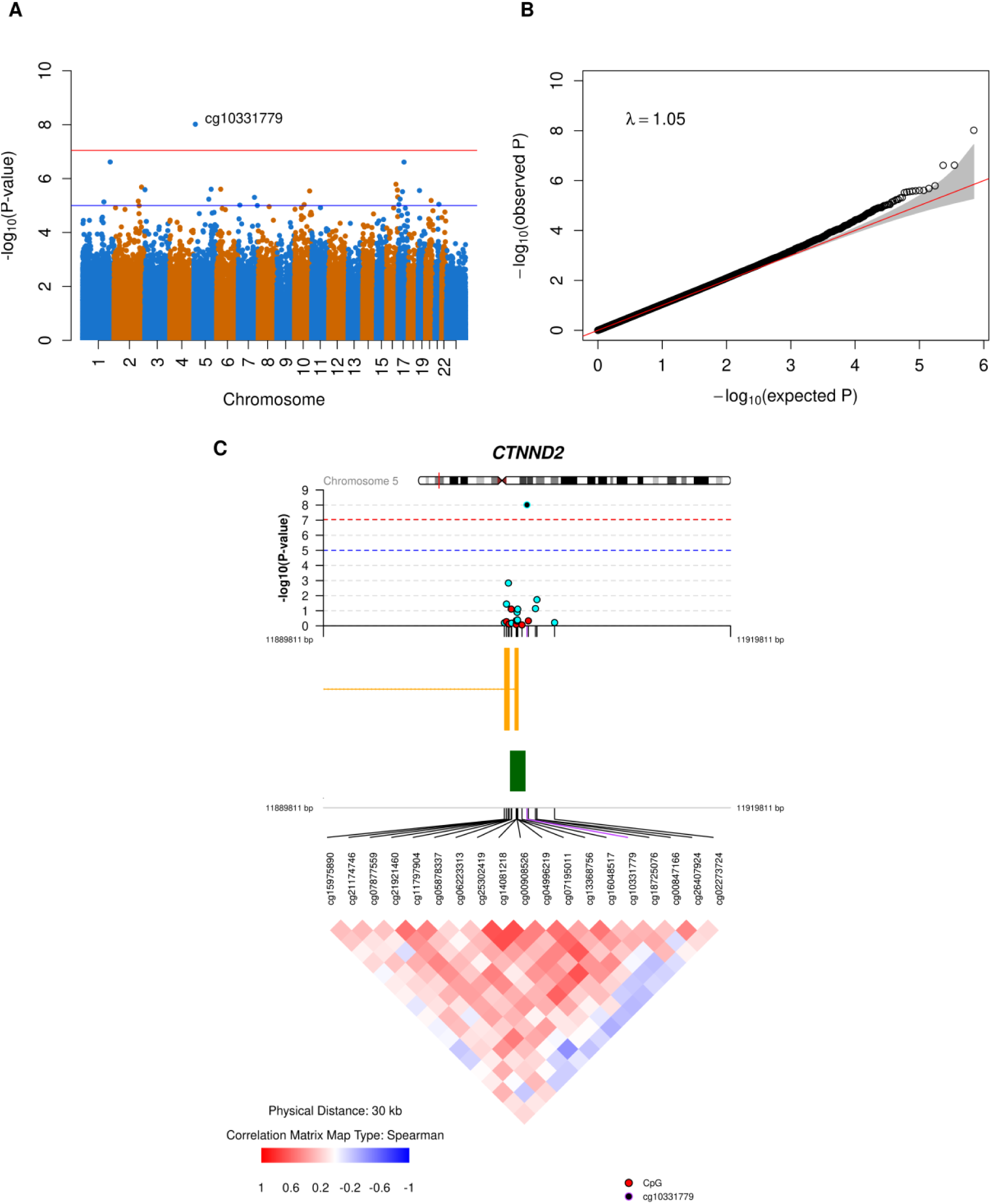
CTH-adjusted EWAS results for processing speed. A. Manhattan plot. The red line represents the epigenome-wide significance threshold with a p-value of 9 × 10^−8^ and the blue line represents the suggestive threshold with a p-value of 1 × 10^−5^. B. QQ plot with 95% confidence interval shaded in grey. The red line represents the reference line where the points should fall under the null hypothesis of no association. C. CoMET plot of a 30 kb region on chromosome 5 centered on cg10331779. The upper panel shows the regional plot of the DMC and its adjacent CpGs. CpGs positively associated with processing speed are displayed in red while CpGs negatively associated with processing speed are displayed in aqua blue. The middle panel shows the gene annotation track (yellow) and CpG island track (green). The lower panel shows the correlation pattern of DNAm beta values for CpGs in this region.

**Figure 3.**
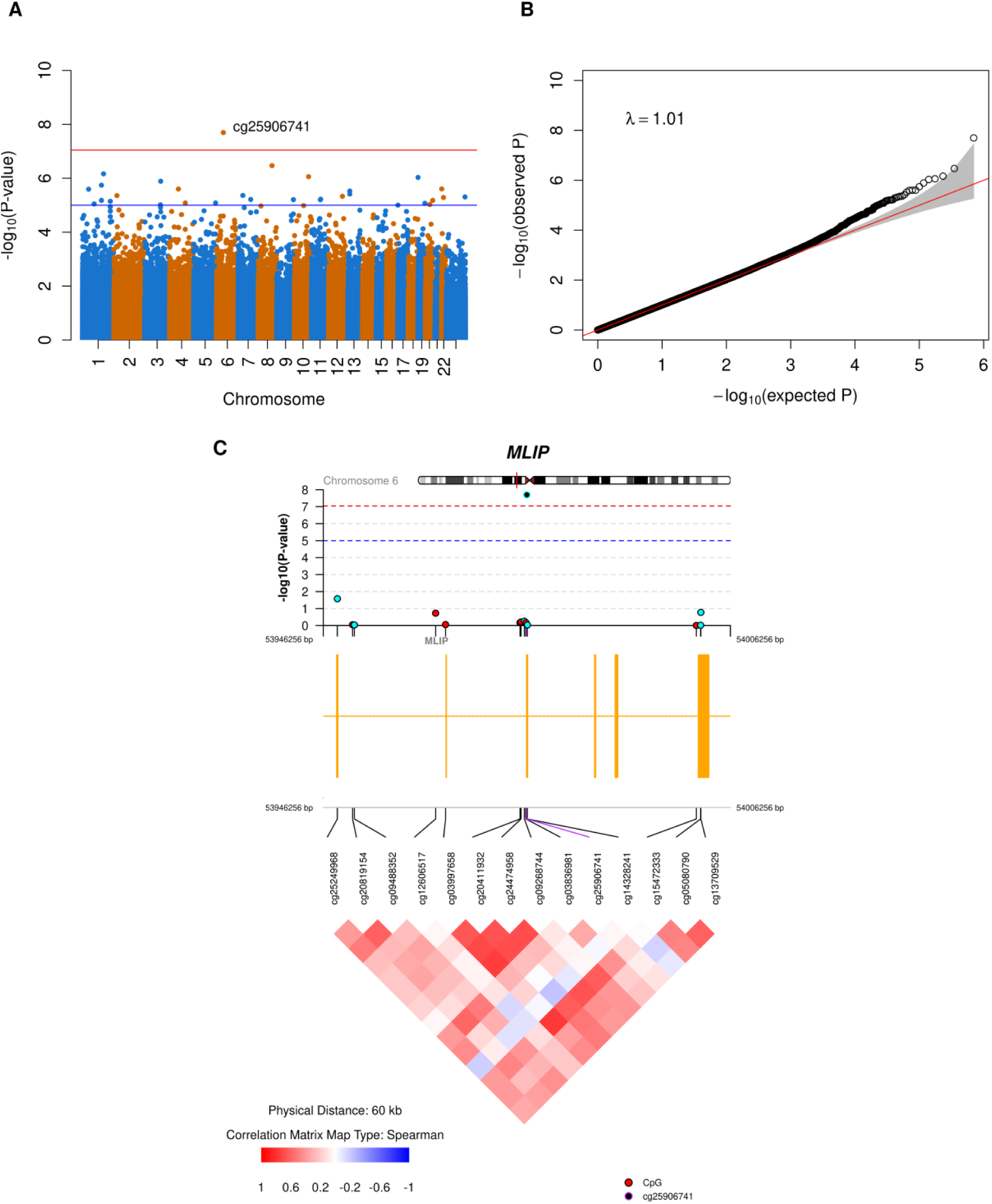
CTH-adjusted EWAS results for PAOFI total scores. A. Manhattan plot. The red line represents the epigenome-wide significance threshold with a p-value of 9 × 10^−8^ and the blue line represents the suggestive threshold with a p-value of 1 × 10^−5^. B. QQ plot with 95% confidence interval shaded in grey. The red line represents the reference line where the points should fall under the null hypothesis of no association. C. CoMET plot of a 60 kb region on chromosome 6 centered on cg25906741. The upper panel shows the regional plot of the DMC and its adjacent CpGs. CpGs positively associated with PAOFI total scores are displayed in red while CpGs negatively associated with PAOFI total scores are displayed in aqua blue. The middle panel shows the gene annotation track and the lower panel shows the correlation pattern of DNAm beta values for CpGs in this region.

**Table 2.** Significant differentially methylated CpG sites of CTH-adjusted EWAS.

| Outcome | CpG name | Chromosome | Position* | Nearest gene | UCSC gene feature category | CpG annotation | Coefficient^ | P-value | FDR Q value |
| --- | --- | --- | --- | --- | --- | --- | --- | --- | --- |
| Processing speed | cg10331779 | 5 | 11904811 | *CTNND2* | TSS1500 | South shore | -0.0369 | 9.65E-09 | 0.0067 |
| PAOFI total scores | cg25906741 | 6 | 53976256 | *MLIP* | 5'UTR;1stExon;Body | Open sea | -0.0027 | 2.01E-08 | 0.0141 |

\*The annotation is based on hg19 genome build
^Coefficients in DNAm $\beta$ -value scale ( $\Delta\beta$ ) were calculated using the formula here<sup>36</sup>

Using the summary statistics from both CTH-unadjusted and CTH-adjusted EWAS, one significant DMR near *RNF26* with a Bonferroni-adjusted p-value less than 0.05 was found for learning and memory without the adjustment of CTH (Table S6). When adjusting for CTH, three DMRs were identified for processing speed, respectively located in/near *SLC6A11, PRKG1/CSTF2T,* and *FAM3B* (Table 3). And DMRs in/near *PI4KB* and *SGCE/PEG10* were detected for mental flexibility. Details of the sensitivity analysis results were described in the supporting information and supplementary tables (Table S23 – Table S24).

**Table 3.** CTH-adjusted differentially methylated regions with adjusted p-value less than 0.05.

| Chromosome | Start position | End position | Number of CpGs | Nearest genes | Adjusted p-value |
| --- | --- | --- | --- | --- | --- |
| Processing speed |  |  |  |  |  |
| 3 | 10857258 | 10857717 | 8 | *SLC6A11* | 3.68E-02 |
| 10 | 53459369 | 53459604 | 6 | *PRKG1; CSTF2T* | 2.59E-02 |
| 21 | 42688191 | 42688675 | 3 | *FAM3B* | 3.19E-03 |
| Mental flexibility |  |  |  |  |  |
| 1 | 151300283 | 151300723 | 9 | *PI4KB* | 5.84E-03 |
| 7 | 94286304 | 94286834 | 18 | *SGCE; PEG10* | 2.82E-04 |

### Post-EWAS functional analysis

Although no significant GO terms or KEGG pathways were identified for any CF phenotype, several canonical pathways and upstream regulators were found from the IPA core analysis (Table S7 – Table S22). For example, the axonal guidance signaling pathway, which is involved in receiving and processing cues to guide the axon to its final destination, was identified as the top one for processing speed and mental flexibility. For PAOFI total scores, the synaptogenesis signaling pathway, which is essential in brain development, was identified as the most significant pathway. Several upstream regulators were shared for most CF phenotypes, including the chemicals beta-estradiol and decitabine, iron channel molecule KCNJ2, growth factor TGFB1, and estrogen receptor ESR1.

No SNPs were found for cg10331779 in the mQTL lookup analyses. One *cis* mQTL rs684940 (chr6:53961513 in *MLIP*) was identified for cg25906741 (p-value=1.65 × 10^−2^^2^, beta=-0.328). However, no associations between this SNP and CF phenotypes were found in the GWAS Catalog (http://www.ebi.ac.uk/gwas/, RRID:SCR_012745)^37^.

Three databases were applied comparing blood and brain DNAm levels. First, the blood brain DNA methylation comparison tool (https://epigenetics.essex.ac.uk/bloodbrain/) showed that cg10331779 in *CTNND2* was significantly correlated with DNAm in the superior temporal gyrus (p-value=0.033, r=0.25), a region involved in language processing, and the mean blood DNAm was higher than mean DNAm in all the four brain regions (prefrontal cortex, entorhinal cortex, superior temporal gyrus, and cerebellum, Figure S24A). Secondly, BECon (https://redgar598.shinyapps.io/BECon/) identified a moderate correlation between blood DNAm and Brodmann area 10, which is linked to higher CFs like decision-making and planning, for cg10331779 (r=0.34, Figure S24B). The third comparison tool, IMAGE-CpG (http://han-lab.org/methylation/default/imageCpG), showed no evidence of correlations between blood and brain DNAm for either cg10331779 or cg25906741.

## DISCUSSION

In this study, we evaluated the relationships between seven objectively measured CF domains from a comprehensive neuropsychological test battery and one subjective CF phenotype and DNAm in early-stage HR+ postmenopausal BC patients. We carried out two types of EWAS with and without the adjustment of CTH, which aided in understanding the role of cell type compositions, followed by DMR analyses. Among the eight CF phenotypes, we identified two DMCs in the CTH-adjusted EWAS, cg10331779 in *CTNND2* with processing speed and cg25906741 in *MLIP* associated with PAOFI total scores, and five DMRs including regions in/near *SLC6A11*, *PRKG1/CSTF2T*, and *FAM3B* for processing speed, and regions in/near *PI4KB* and *SGCE/PEG10* for mental flexibility. Post-EWAS analyses were performed to identify the biological processes and pathways associated with CF.

The top DMC cg10331779 in the CTH-adjusted processing speed EWAS, with p-value=9.65 × 10^−9^, maps to the promoter region in *CTNND2*. Although in the sensitivity analysis using only participants who had not received ET this CpG did not reach genome-wide significance (p-value=1.14 × 10^−4^), their coefficients were similar with –0.0369 in the main analysis and – 0.0398 in the sensitivity analysis (Figure S14). This potentially indicates that ET may not be driving the variation in methylation at cg10331779 associated with processing speed. Methylation of cg10331779 was also associated with poorer mental flexibility at a nominal significance level (p-value=0.018, Table S3). Further searching found that DNAm levels of cg10331779’s of blood correlated with those in the superior temporal gyrus, an area involved in language processing, and Brodmann area 10, which contributes to high-level CF including decision-making and prospective memory^38–40^. *CTNND2,* which encodes an adhesive junction-associated protein delta catenin and is mainly expressed in the brain, has been widely studied in neurocognitive function^41,42^. Deletion of chromosome 5p where *CTNND2* is located can cause Cri-du-chat syndrome which is characterized by a high-pitched cat-like cry, intellectual disability, and poor development^43,44^. Earlier studies showed that polymorphisms in *CTNND2* may be associated with cognitive ability^45–47^. For instance, rs61749834 was a suggestive SNP of processing speed, the CF domain where differential methylation in *CTNND2* was detected in the current study, for teenagers between 10 and 17 years of age^45^. SNPs rs62337555 and rs12519314 were associated with general cognitive performance in a multivariate GWAS when jointly analyzed with education-related variables and personality phenotype, respectively^46,47^. Furthermore, genetic variations in *CTNND2* were shown to be linked with a variety of neurodevelopmental disorders, including autism, attention deficit hyperactivity disorder (ADHD), depression, Alzheimer’s disease (AD), and myopia^48–55^. *CTNND2* is also known as a modulator of the Wnt/β-catenin signaling pathway which is associated with several diseases and disorders, including cancer and neurodegenerative diseases^56–58^. Our results from the IPA core analysis showed that a transcription regulator SOX2, which also regulates the Wnt/β-catenin signaling pathway, was identified as an upstream regulator for processing speed, playing an important role in nervous system development, neurodevelopmental disorders, and adult brain function^59,60^.

DNAm variation at cg25906741 in *MLIP* was significantly associated with subjective PAOFI total scores in the main CTH-adjusted EWAS (p-value=2.01 × 10^−8^) and it was a suggestive DMC in the sensitivity analysis (p-value=5.18 × 10^−6^). In previous research, *MLIP* was found to be related to AD development and ADHD^61,62^. While *MLIP* is known to interact with lamin A/C (LMNA) associated with muscle-related disorders, studies on *MLIP* in CF and neurodevelopment are limited^63–67^. Cluett et al. showed that polymorphism in the *LMNA* gene was associated with CF, as measured by the Mini-Mental State Examination, in older adults^68^. In addition, mRNA levels of LMNA were increased in the hippocampus for patients with late-stage AD^69^. It is possible that the biological mechanisms of CF impairment in BC patients can partially overlap with AD pathology with the interaction between *MLIP* and *LMNA*.

We compared the associations of these two significant DMCs identified in our current study with a recently published EWAS study in the Generation Scotland cohort. McCartney and colleagues evaluated the relationship between DNAm and several CF measurements, including digit symbol tests, logical memory, verbal fluency, vocabulary, general fluid CF, and general CF^70^. Neither DMC showed significant associations with any CF phenotypes measured in the McCartney et al study (Table S4, Figure S25).

In our study, we also noticed the potential impact of estrogen on CF for HR+ BC patients when identifying the upstream regulators using IPA. Beta-estradiol, a steroid estrogen hormone, was observed as an upstream regulator for all eight cognitive function phenotypes in both the main and the sensitivity analyses. It is converted from testosterone by aromatase and binds to the estrogen receptors (ER) to regulate ER target genes, and it plays an important part in HR+ BC by increasing the risks of cancer cell growth and migration^71,72^. Beta-estradiol is associated with CF and may be involved in the promotion of memory^73^. Therefore, based on previous evidence and our results, we hypothesize that ET, as a popular therapy to treat HR+ BC, may disrupt the synthesis or the activity of beta-estradiol that impacts CF by regulating downstream CF-related genes. Even in patients free of ET, levels of beta-estradiol also regulate CF (results not shown). However, future studies will be required to demonstrate this relationship.

We identified several CF-associated DMRs mapping to genes that maintain normal cognition and brain functions. Among these, *SLC6A11*, which encodes a GABA transporter protein, is associated with many neurological disorders including epilepsy and intellectual disability^74,75^. Additionally, *PRKG1* and *CSTF2T* may be associated with learning ability in previous reports^76,77^, while *SGCE* and *PEG10*, both imprinted genes, are connected to psychiatric disorders and tumor progression, respectively^78,79^. Our results provide additional evidence that variations in DNAm across these genes may be associated with variation in CF in BC patients.

Our study has several potential limitations. First, due to the small sample size, this study may miss true DMCs associated with CF and could produce false positives. Second, our discoveries have not been validated in independent cohorts. Larger validation cohorts can help determine whether the results are robust. Finally, the heterogeneity of ET status should not be neglected. Because of the pandemic, the original enrollment criteria were changed and so some participants had already started ET when their blood sample was collected. We addressed this by conducting a sensitivity analysis where only participants who were free of ET at blood sample collection were included. By comparing the main and sensitivity analyses, the significant DMCs were different. The observed differences were not surprising given the smaller sample size used in the sensitivity analyses, taking the potential influence of the winner’s curse into account^80^. Although the p-values differed, the effect sizes of the significant and suggestive DMCs were similar between the two analyses (Figure S12-S19). In addition, since EPICC participants were treated with ET after enrollment (if not before) and followed up for six months, we plan to conduct subsequent studies to enhance the understanding of how ET impacts DNAm signatures and CF.

In conclusion, our study evaluated the relationship between DNAm variations and CF phenotypes in early-stage HR+ BC patients and identified several CpGs and DMRs that may enhance the understanding of the biological mechanisms behind CF performance in this population. Further validation will be necessary to verify our discoveries.

## AUTHOR CONTRIBUTIONS

Shuwei Liu: Data curation, formal analysis, investigation, methodology, visualization, writing – original draft preparation, writing – review & editing

Dongjing Liu: Investigation, methodology, writing – review & editing

Catherine Bender: Conceptualization, funding acquisition, writing – review & editing

Kirk Erickson: Conceptualization, funding acquisition, writing – review & editing

Susan Sereika: Data curation, methodology, writing – review & editing

John Shaffer: Conceptualization, methodology, funding acquisition, supervision, writing – review & editing

Daniel Weeks: Conceptualization, investigation, formal analysis, methodology, resources, funding acquisition, supervision, writing – review & editing

Yvette Conley: Conceptualization, methodology, resources, funding acquisition, supervision, project administration, writing – review & editing

## DATA AVAILABILITY STATEMENT

The EPICC data supporting the findings of this study are available upon reasonable request from the corresponding author.

## IRB STATEMENT

This study was reviewed and approved by the Institutional Review Board (IRB) of the University of Pittsburgh (STUDY19080223 and STUDY19010174). All participants provided written consent forms at enrollment.

## Condensed abstract

Variations of DNA methylation are associated with cognitive function in early-stage HR+ breast cancer patients. Beta-estradiol was identified as a common upstream regulator for all the cognitive function phenotypes.

## Acknowledgments

This study is a part of the Exercise Program for Cognition in Cancer (EPICC) randomized clinical trial and was funded by National Institute of Health grants R01CA221882 (Conley, Bender, Erickson) and R01CA196762 (Bender, Erickson). We express gratitude to all the participants and their families.

