## Supporting information for "Associations between DNA methylation and cognitive function in early-stage hormone receptor-positive breast cancer patients"

**for**

#### Study workflow

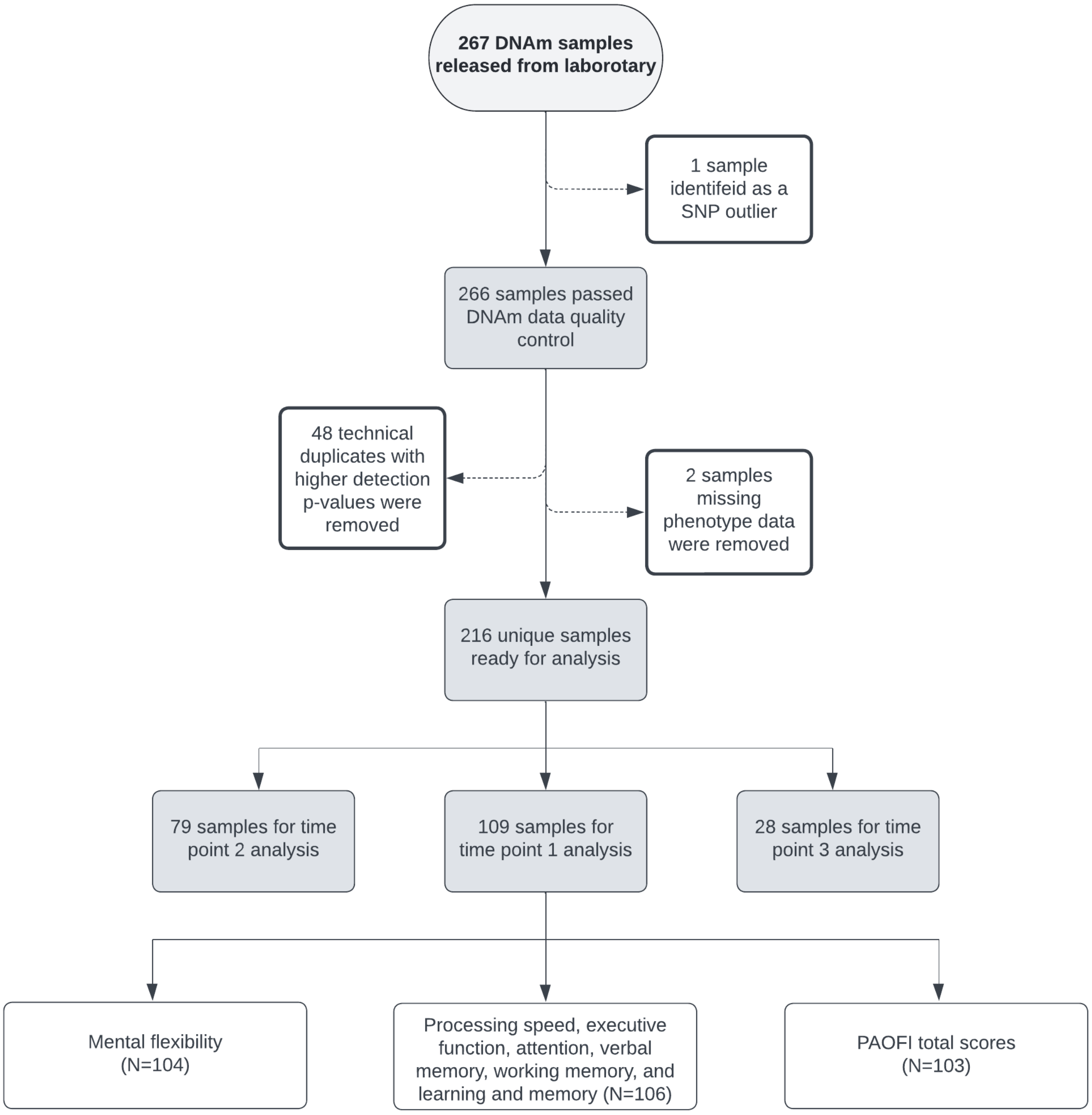

**Figure S1. Overview of the data collection, quality control of three time points**

#### DNAm data quality control

##### Sample-level quality check

Our work is a part of a longitudinal study, although just baseline analyses were carried out in the current manuscript. DNAm of three time points were assayed in three batches, each consisting of samples from different time points, and the quality control was performed together.

We first examined the density of DNAm β values, mean detection p-values, bisulfite conversion intensity, and percentage of low-quality CpGs in each sample to have an overview of the sample-level quality[1–3]. Samples with abnormal distribution of β values, high mean detection p-values, low bisulfite conversion intensity, or high percentage of low-quality CpGs should be removed.

As shown in the β value density plot below (Figure S2), DNAm β values across samples displayed two peaks and were multimodal distributed.

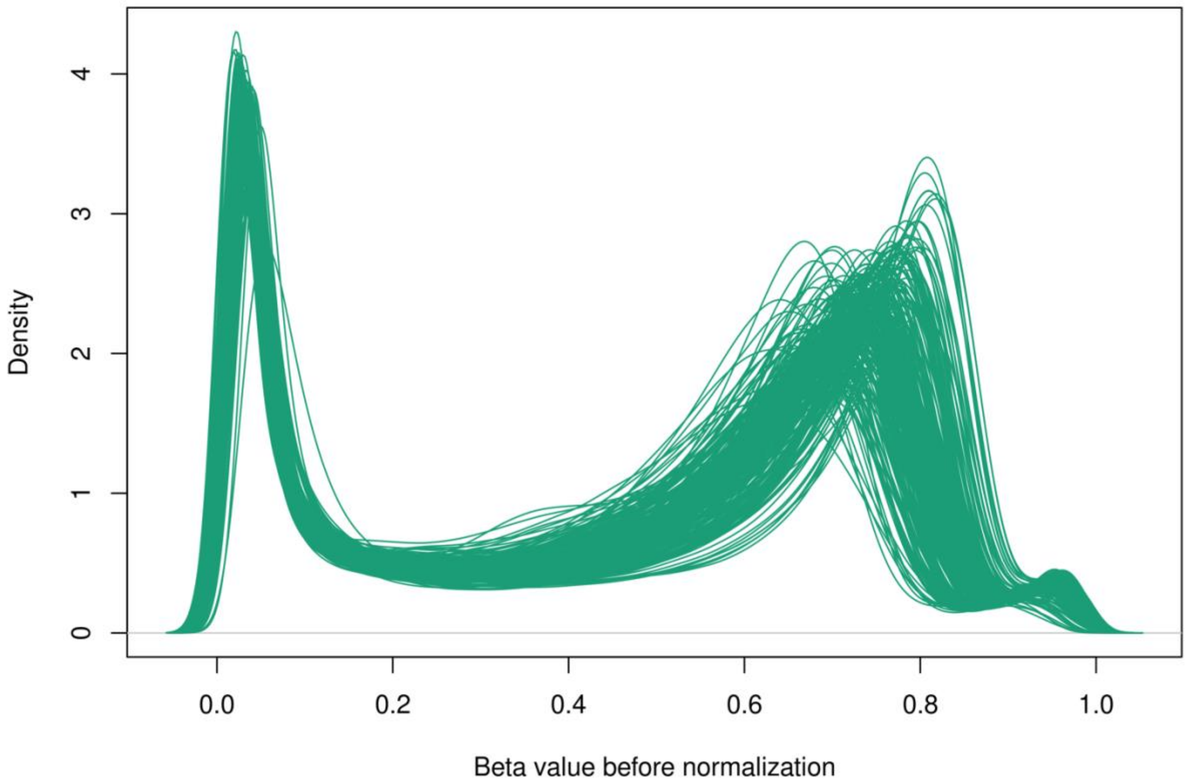

**Figure S2. Density plot of the raw DNAm β values**

With the threshold of 0.01 (red line), no samples were removed due to having high mean detection p-values (Figure S3).

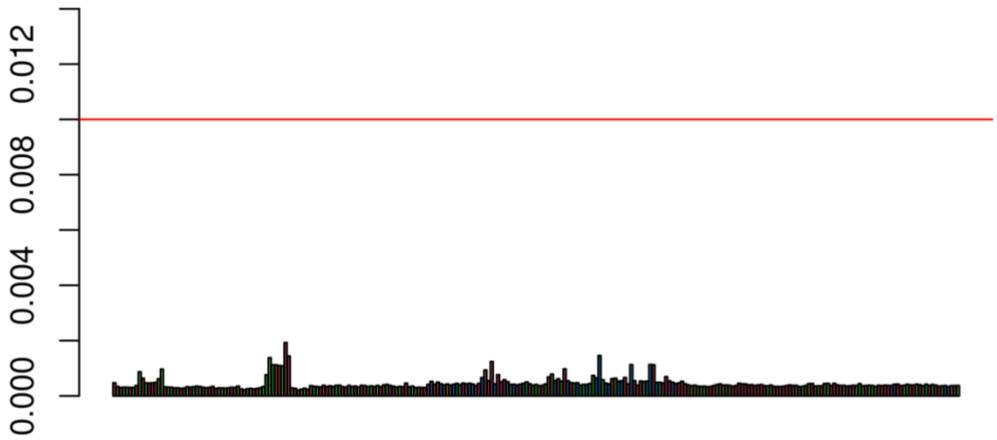

**Figure S3. Mean detection p-values for each sample**

ENmix identified 6 samples with a higher percentage of low-quality CpGs (>0.01). However, as shown in the plot below, these samples were not removed in this step as they were very close to the 0.01 threshold (Figure S4).

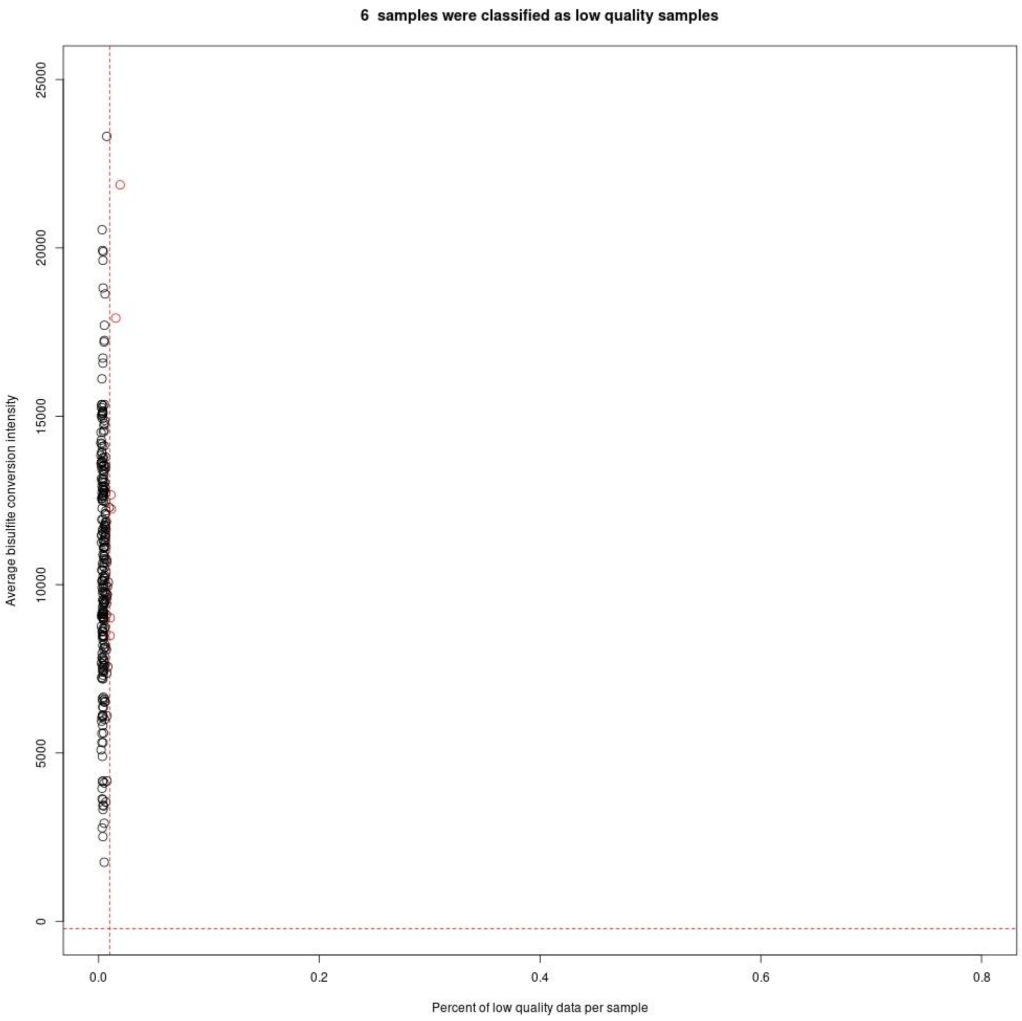

**Figure S4. Sample quality plot based on percentage of low-quality CpGs and bisulfite conversion intensity**

In addition to the common sample-level quality checks mentioned above, we also evaluated the distribution of β values for SNP probes[4]. Usually, β values of the SNP probes should cluster in one of the three categories (AA, AB, BB). Outliers may indicate contaminated or failed samples. In our case, one sample was identified as an outlier and thus got removed (Figure S5).

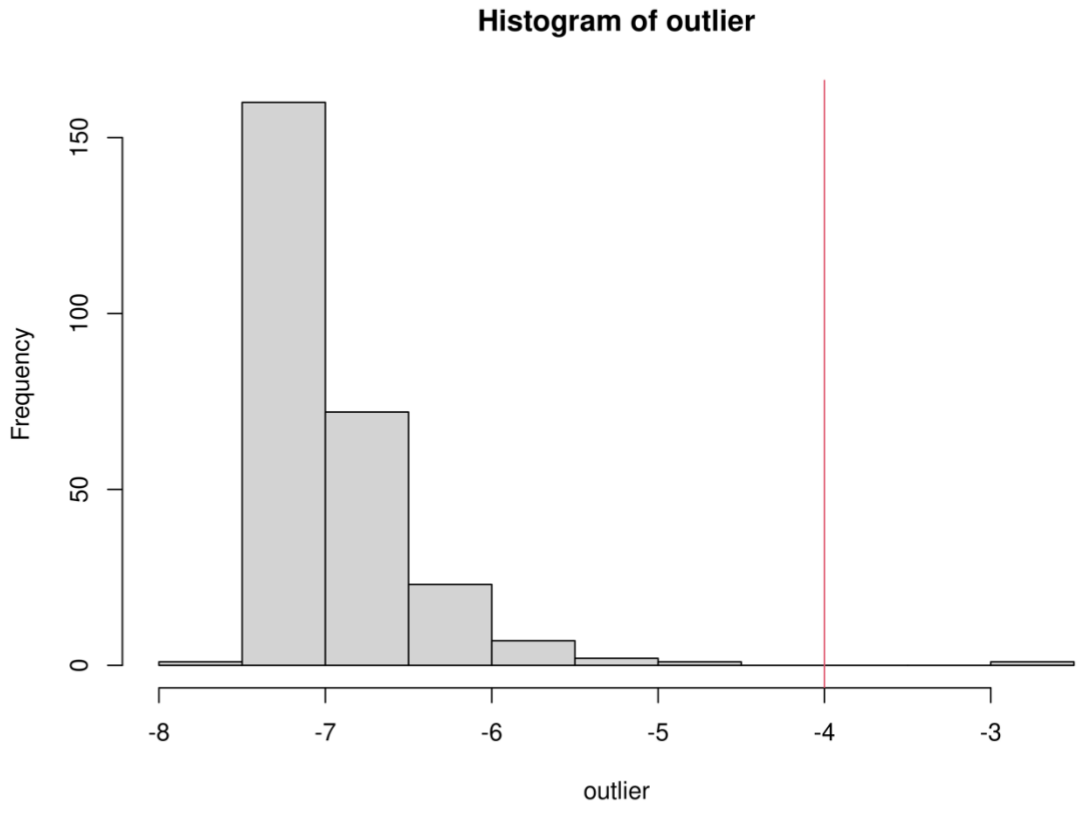

**Figure S5. Histogram of SNP outliers**

Evaluate the average log odds of deviating from the expected genotypes across all the SNPs

**Normalization and removal of low-quality probes**

Next, functional normalization with dye bias and background correction was performed by minfi[1,2].

CpGs were removed if they were (1) probes with SNPs at CpG or single base extension (SBE) sites, (2) cross-reactive probes, (3) probes on the Y chromosome, and (4) "bad" probes identified by ENmix as shown in Table S1[3].

**Table S1. Number of CpGs removed in each quality control step**

| Quality control step | Number of CpGs | Number of removed CpGs |
|---|---|---|
| **Raw** | 865,859 | NA |
| **Probes with SNPs** | 835,424 | 30,435 |
| **Cross-reactive probes** | 708,328 | 127,096 |
| **Y chromosome probes** | 707,949 | 379 |
| **ENmix bad probes*** | 700,779 | 7,170 |

* ENmix bad probes include CpG probes whose percentage of low-quality CpG sites across all samples was greater than 0.05, whose detection p-values were greater than 0.01, or whose number of beads was less than 3.

After normalization and removal of bad probes, the quality of DNAm data was improved as the DNAm β values were bimodally distributed (Figure S6) and showed better clustering (Figure S7-S10).

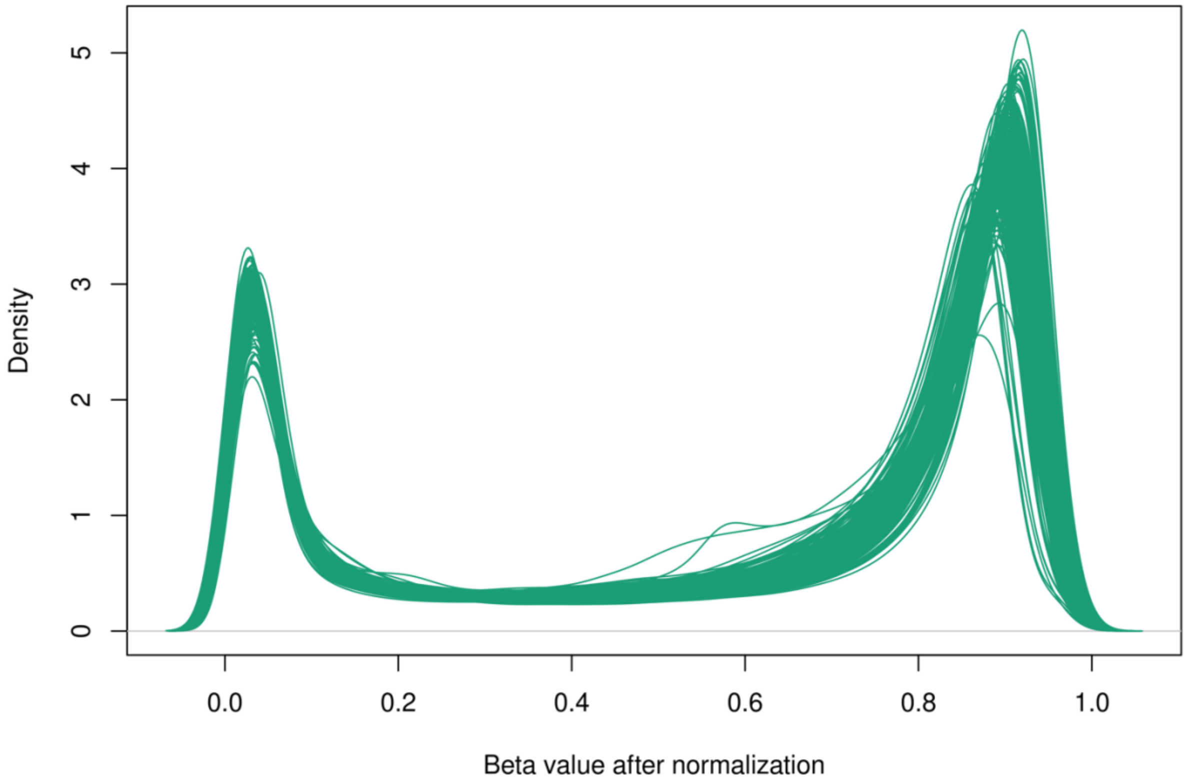

**Figure S6. Density plot of the normalized DNAm β values**

Taking advantage of technical duplicates within or across batches, we can also evaluate the overall performance of quality control procedures by comparing the number of clustering pairs before and after normalization under the assumption that technical duplicates should cluster with each other (Figure S6-S9). Technical duplicates were well-clustered with each other after quality control procedures only 3 pairs were not aligned, compared with 6 pairs for the raw data for both time points.

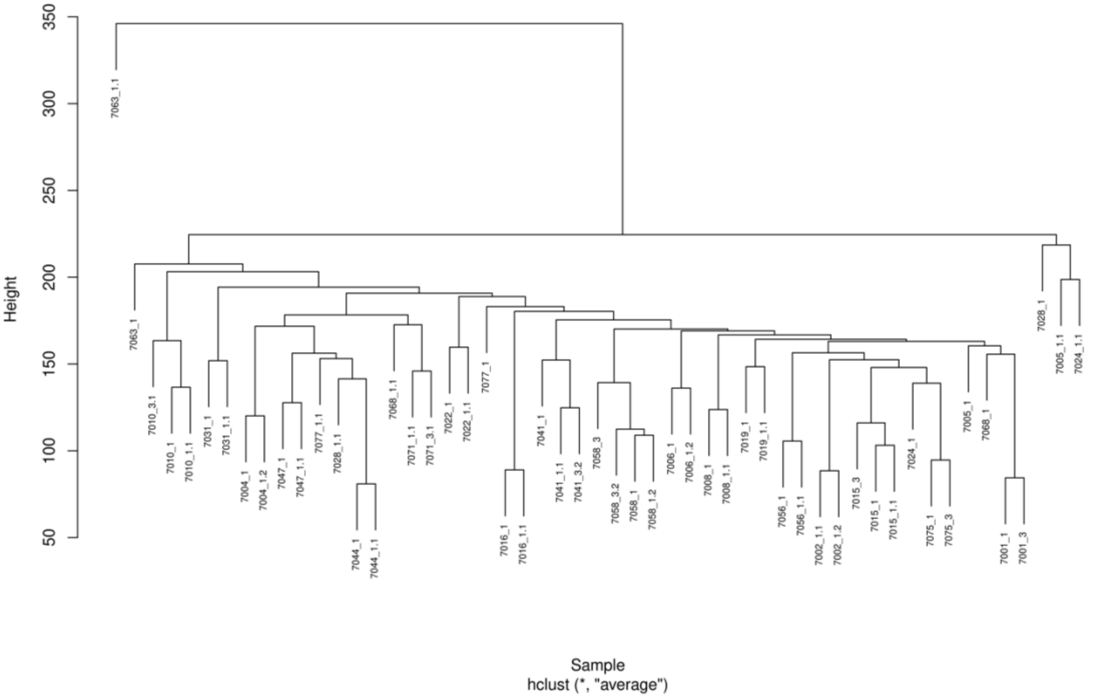

**Figure S7. Clustering plot of technical duplicates based on raw β values (time point 1)**

Notes on IDs: IDs end with “1” – samples measured in the first batch, IDs end with “1.1” – samples measured in the second batch, IDs end with “1.2” – samples measured in the third batch, IDs end with “3” – technical replicates measured in the first batch, IDs end with “3.1” – technical replicates measured in the second batch, IDs end with “3.2” – technical replicates measured in the third batch.

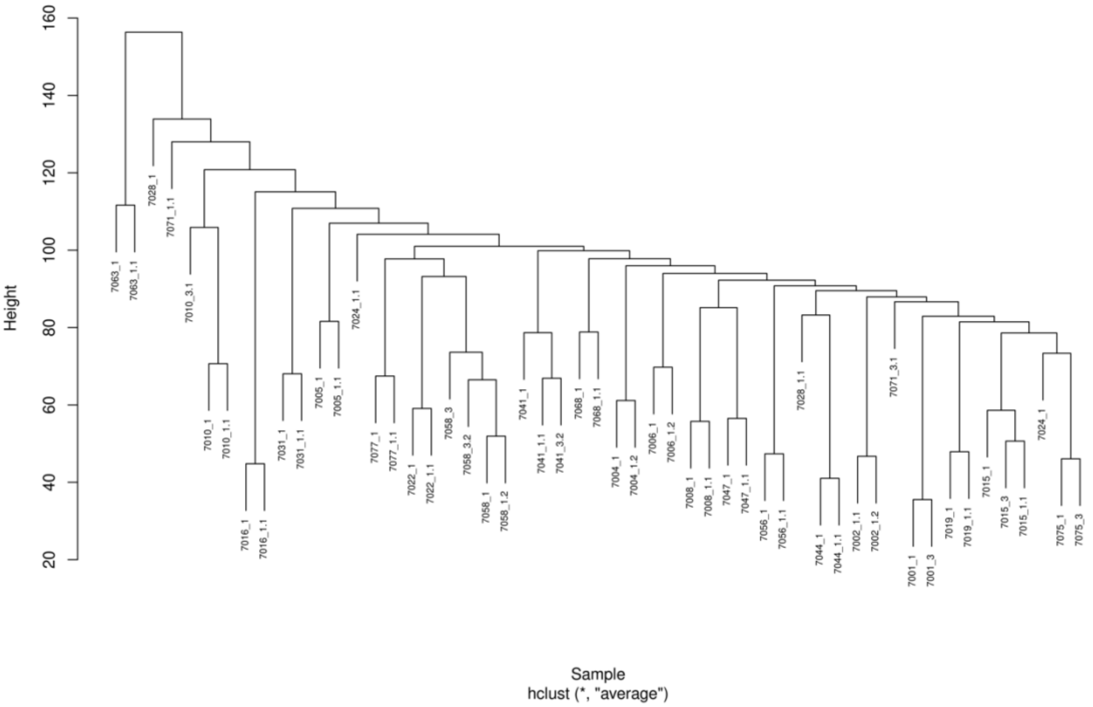

**Figure S8. Clustering plot of technical duplicates based on QCed β values (time point 1)**

Notes on IDs: IDs end with "1" – samples measured in the first batch, IDs end with "1.1" – samples measured in the second batch, IDs end with "1.2" – samples measured in the third batch, IDs end with "3" – technical replicates measured in the first batch, IDs end with "3.1" – technical replicates measured in the second batch, IDs end with "3.2" – technical replicates measured in the third batch.

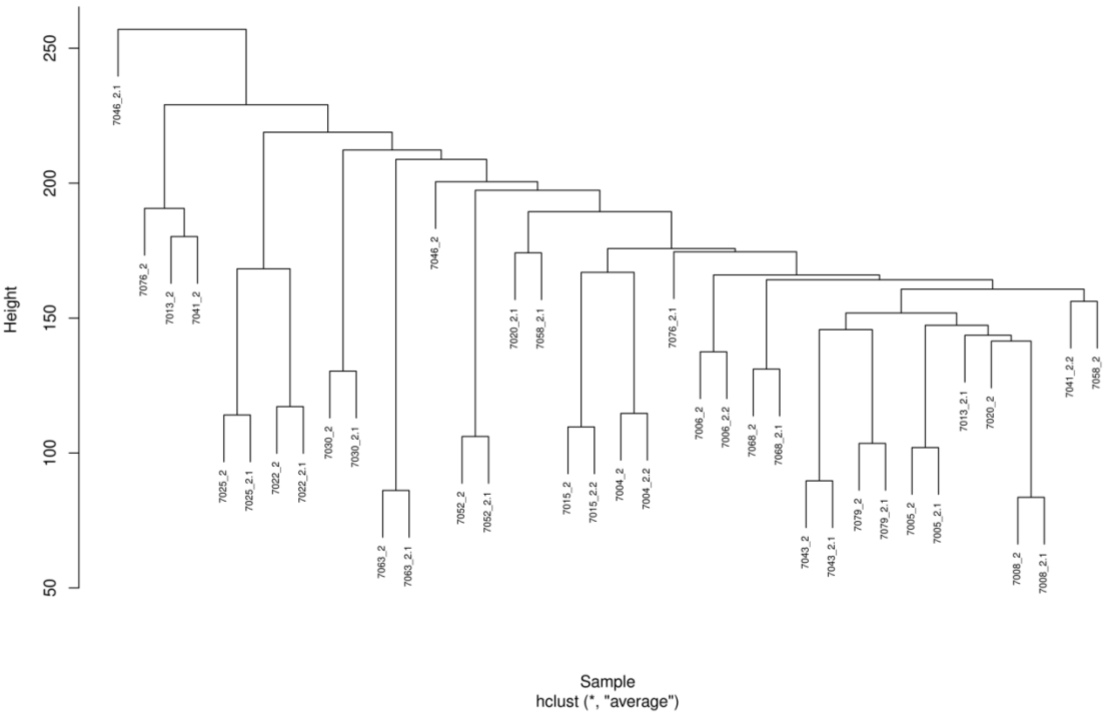

**Figure S9. Clustering plot of technical duplicates based on raw β values (time point 2)**

Notes on IDs: IDs end with “2” – samples measured in the first batch, IDs end with “2.1” – samples measured in the second batch, IDs end with “2.2” – samples measured in the third batch.

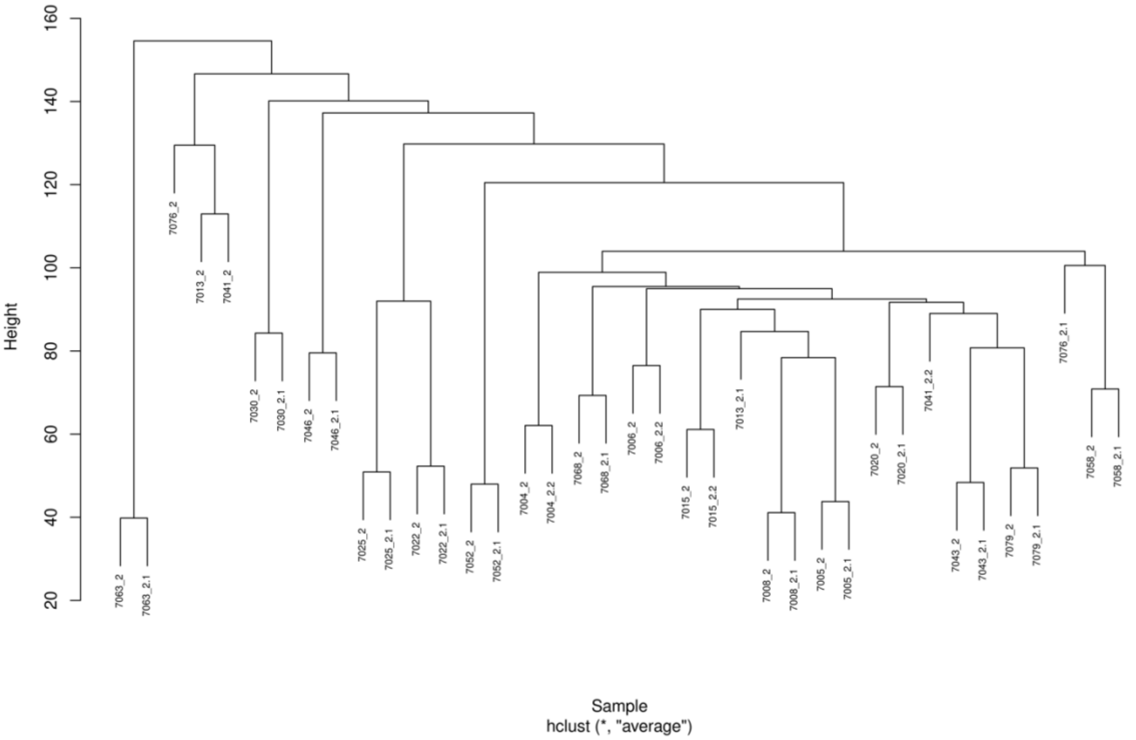

**Figure S10. Clustering plot of technical duplicates based on QCed β values (time point 2)**

Notes on IDs: IDs end with “2” – samples measured in the first batch, IDs end with “2.1” – samples measured in the second batch, IDs end with “2.2” – samples measured in the third batch.

#### Results

#### Sample characteristics

**Table S2. Sample characteristics for continuous variables (A) and categorical variables (B) in samples with DNAm measured and in the EPICC samples**

A. Continuous variables

| | EWAS Sample (N=109) | | | | | | | | EPICC Sample (N=158) | | | | | | | |
|---|---|---|---|---|---|---|---|---|---|---|---|---|---|---|---|---|
| Characteristic | Missing | Min | Q1 | Median | Mean | Q3 | Max | Distribution | Missing | Min | Q1 | Median | Mean | Q3 | Max | Distribution |
| Age | 0 | 24.00 | 58.00 | 64.00 | 62.35 | 67.00 | 78.00 | | 0 | 24.00 | 58.00 | 63.00 | 62.14 | 67.75 | 78.00 | |
| Education in years | 0 | 12.00 | 14.00 | 16.00 | 16.21 | 18.00 | 23.00 | | 1 | 12.00 | 14.00 | 16.00 | 16.00 | 18.00 | 26.00 | |
| Verbal IQ scores | 0 | 84.20 | 109.12 | 112.68 | 112.59 | 118.02 | 125.14 | | 0 | 84.20 | 108.45 | 112.68 | 112.31 | 117.13 | 125.14 | |
| PAOFI total scores | 6 | 0.00 | 7.00 | 15.00 | 20.11 | 27.44 | 83.00 | | 6 | 0.00 | 8.00 | 18.00 | 21.34 | 29.49 | 84.00 | |
| Cognitive function domains (z scores) | | | | | | | | | | | | | | | | |
| Executive function | 3 | -1.96 | -0.07 | 0.32 | 0.23 | 0.67 | 1.15 | | 5 | -1.96 | -0.04 | 0.32 | 0.26 | 0.68 | 1.38 | |
| Attention | 3 | -2.17 | -0.60 | -0.11 | -0.21 | 0.26 | 1.01 | | 5 | -2.98 | -0.61 | -0.12 | -0.25 | 0.25 | 1.01 | |
| Mental flexibility | 5 | -3.03 | -0.19 | 0.32 | 0.11 | 0.65 | 1.06 | | 7 | -3.86 | -0.22 | 0.26 | 0.01 | 0.62 | 1.06 | |
| Learning and memory | 3 | -2.09 | -0.60 | -0.12 | -0.13 | 0.44 | 1.26 | | 5 | -2.23 | -0.64 | -0.12 | -0.15 | 0.39 | 1.26 | |
| Working memory | 3 | -1.63 | -1.03 | -0.52 | -0.40 | -0.03 | 1.66 | | 6 | -1.63 | -1.02 | -0.49 | -0.39 | -0.02 | 1.66 | |
| Verbal memory | 3 | -2.04 | -1.08 | -0.52 | -0.38 | 0.20 | 2.29 | | 5 | -2.28 | -1.08 | -0.52 | -0.40 | 0.20 | 2.29 | |
| Processing speed | 3 | -2.29 | -0.25 | 0.21 | 0.09 | 0.51 | 1.34 | | 5 | -3.01 | -0.41 | 0.13 | -0.02 | 0.52 | 1.34 | |

B. Categorical variables

| Characteristic | N=109<br>EWAS Sample, n (%) | N=158<br>EPICC Sample, n (%) |
|---|---|---|
| Race | | |
| Asian | 1 (0.9%) | 1 (0.6%) |
| Black or African American | 8 (7.3%) | 10 (6.3%) |
| Other | 1 (0.9%) | 2 (1.3%) |
| White | 99 (91%) | 145 (92%) |
| Ethnicity | | |
| Hiapanic or Latino | 1 (0.9%) | 2 (1.3%) |
| Non-Hispanic or non-latino | 107 (99%) | 155 (99%) |
| Missing | 1 | 1 |
| Tumor stage | | |
| DCIS | 16 (15%) | 22 (14%) |
| stage 1 | 68 (62%) | 102 (65%) |
| stage 2a | 15 (14%) | 21 (13%) |
| stage 2b | 5 (4.6%) | 7 (4.4%) |
| stage 3a | 5 (4.6%) | 6 (3.8%) |
| Had chemotherapy before | | |
| Yes | 18 (17%) | 28 (18%) |

#### Executive function

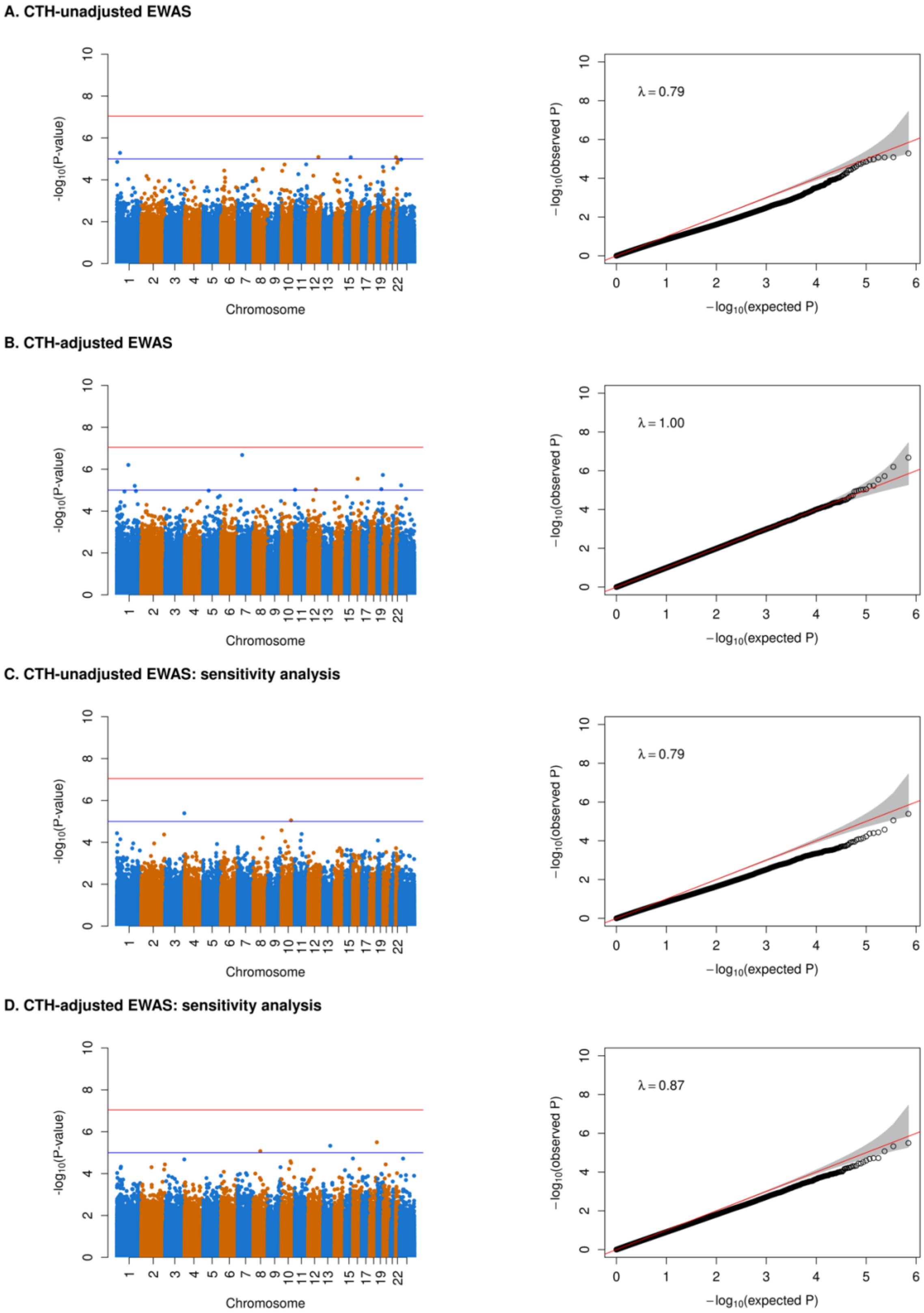

**Figure S11a. EWAS results for executive function**

In the Manhattan plots, the red line indicates the epigenome-wide significance threshold of $9 \times 10^{-8}$ and the blue line indicates the suggestive threshold of $1 \times 10^{-5}$. The $\lambda$ values in the QQ plots were estimated using *estlambda* function in GenABEL package.

### Attention

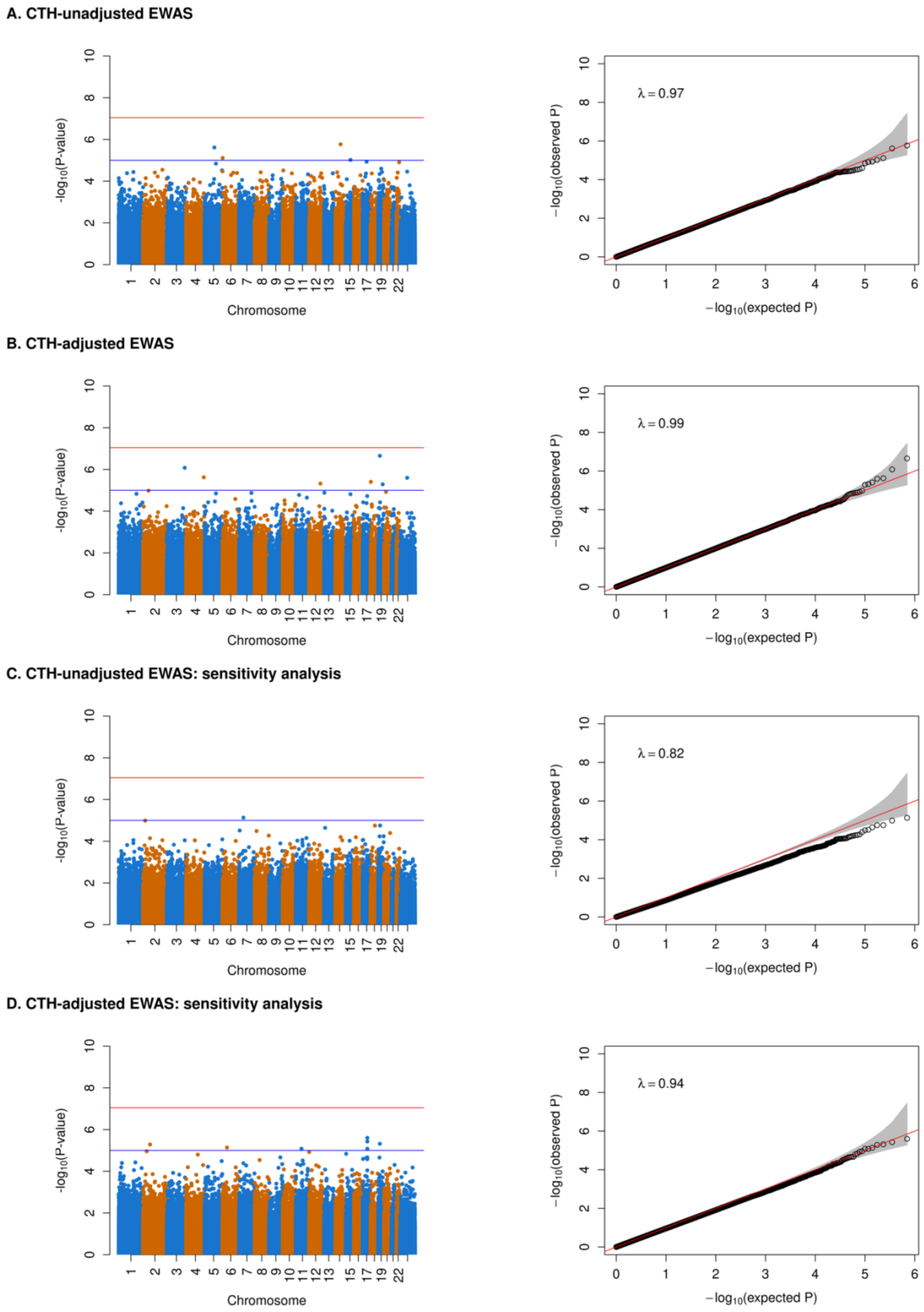

**Figure S11b. EWAS results for attention**

**Mental flexibility**

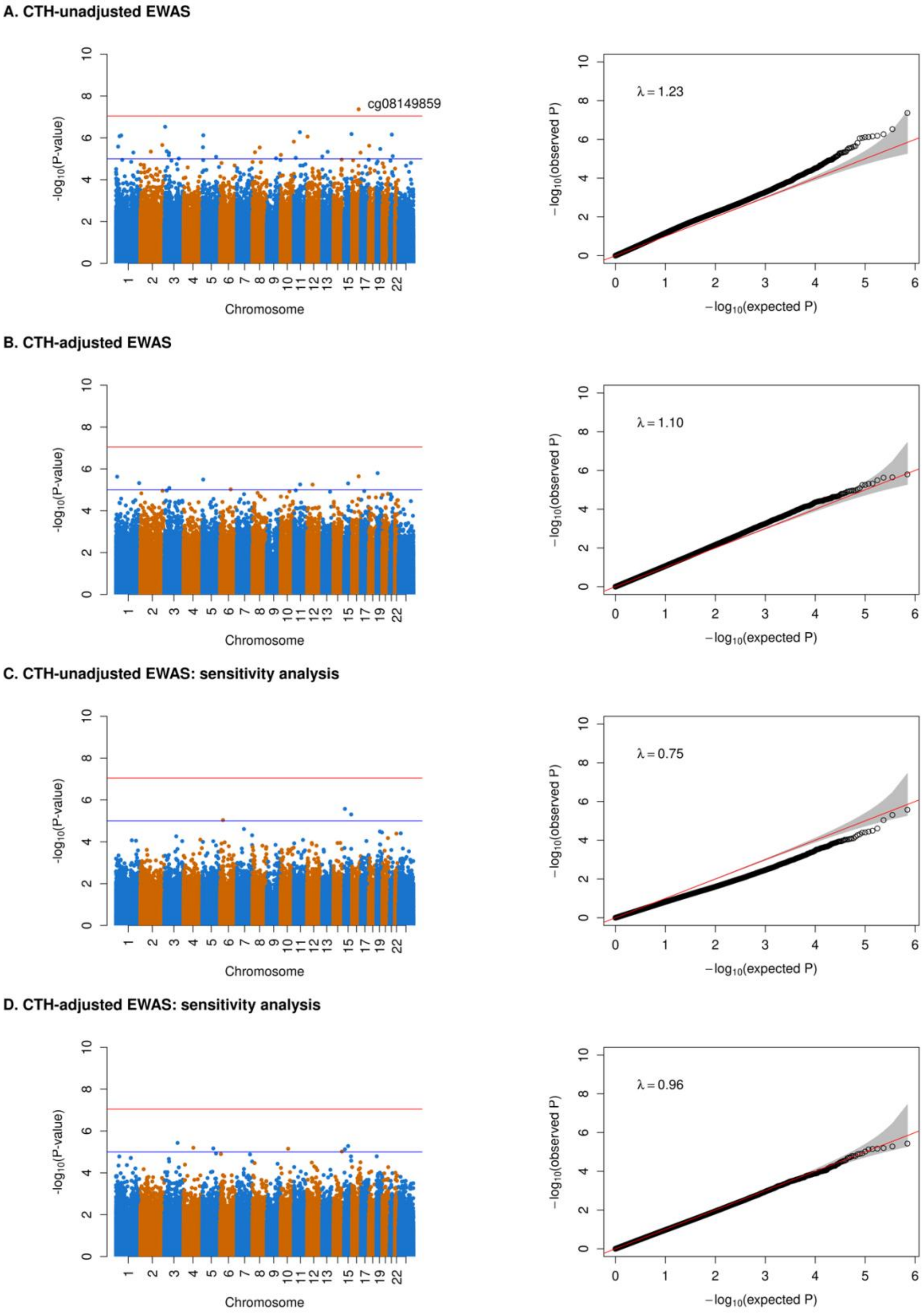

**Figure S11c. EWAS results for mental flexibility**

#### Learning and memory

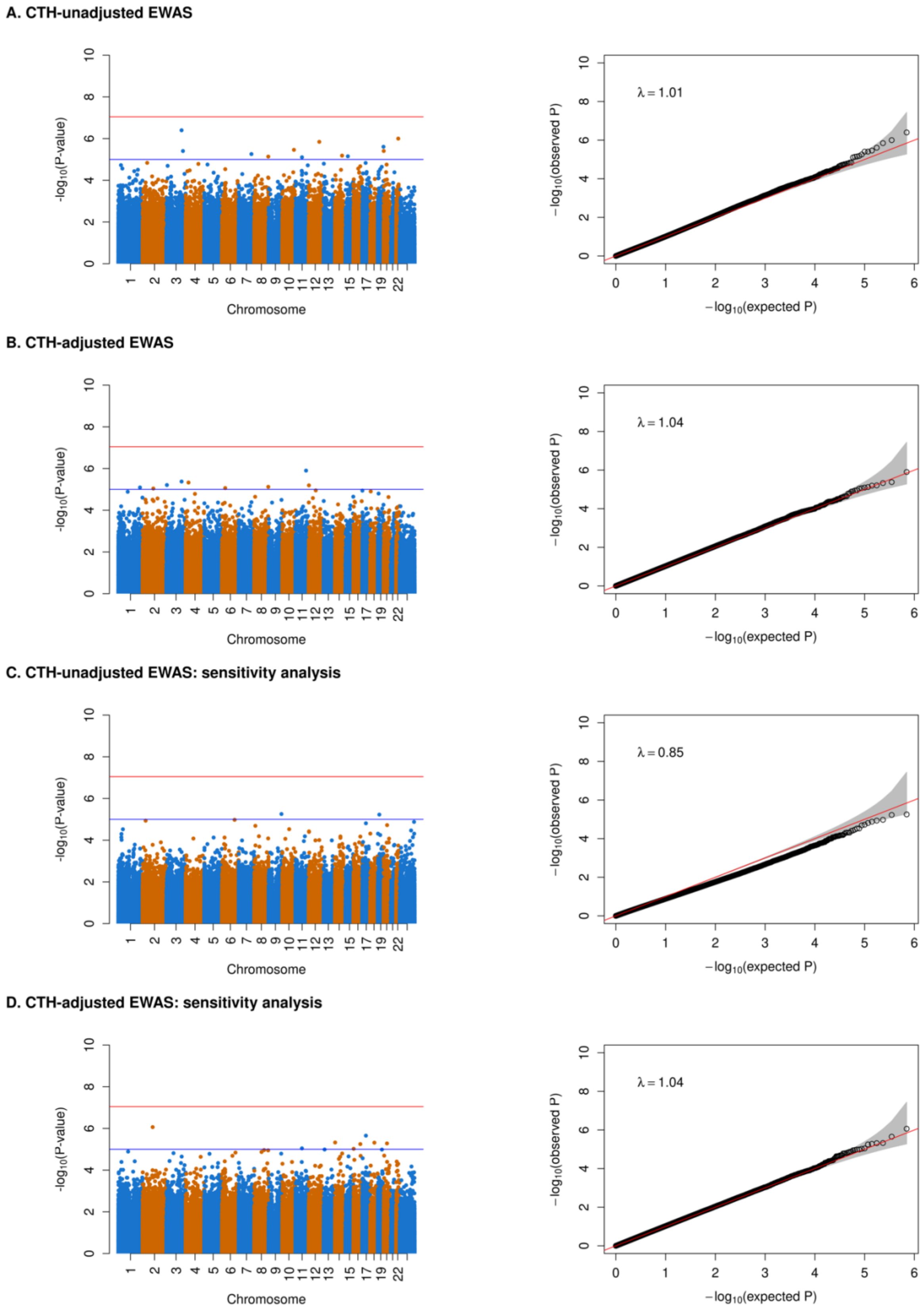

**Figure S11d. EWAS results for learning and memory**

#### Verbal memory

**A. CTH-unadjusted EWAS**

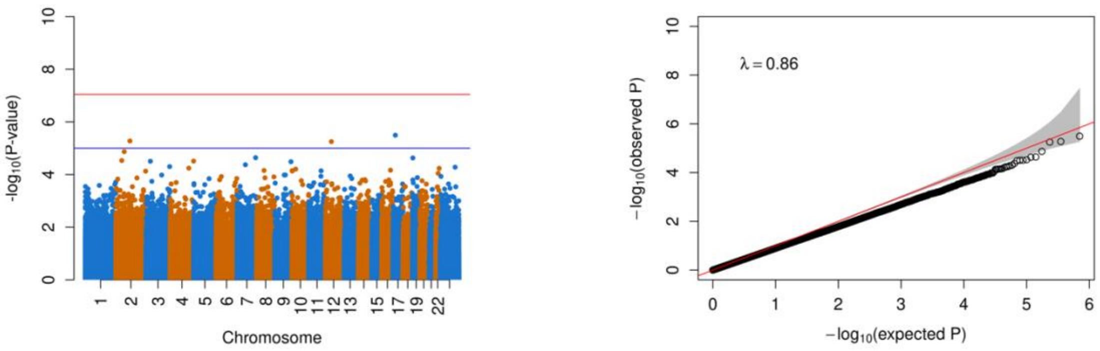

**B. CTH-adjusted EWAS**

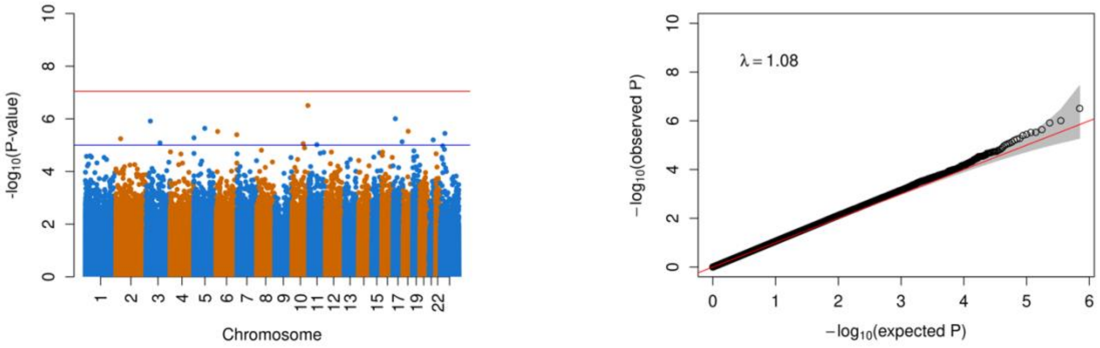

**C. CTH-unadjusted EWAS: sensitivity analysis**

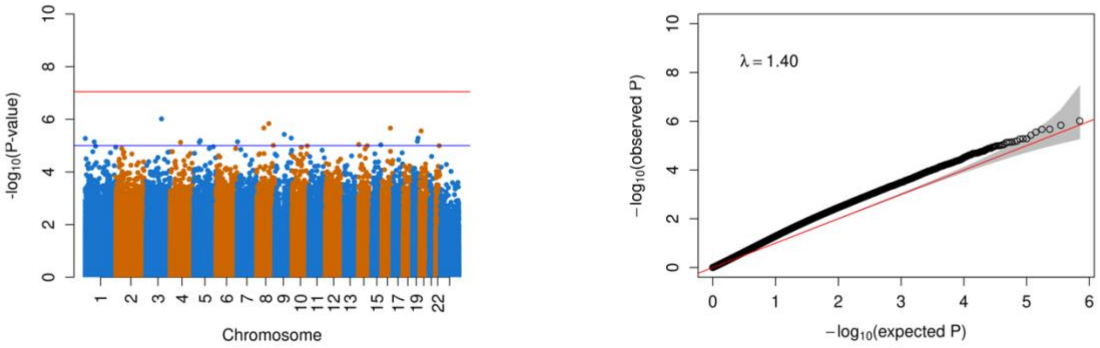

**D. CTH-adjusted EWAS: sensitivity analysis**

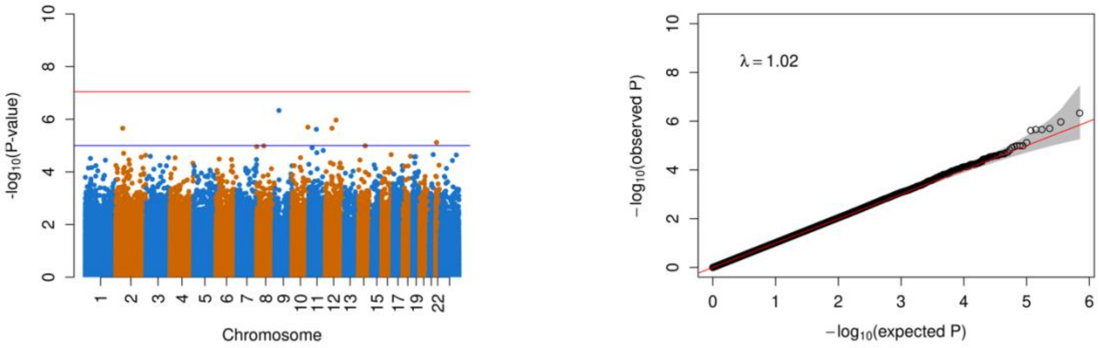

**Figure S11e. EWAS results for verbal memory**

#### Working memory

**A. CTH-unadjusted EWAS**

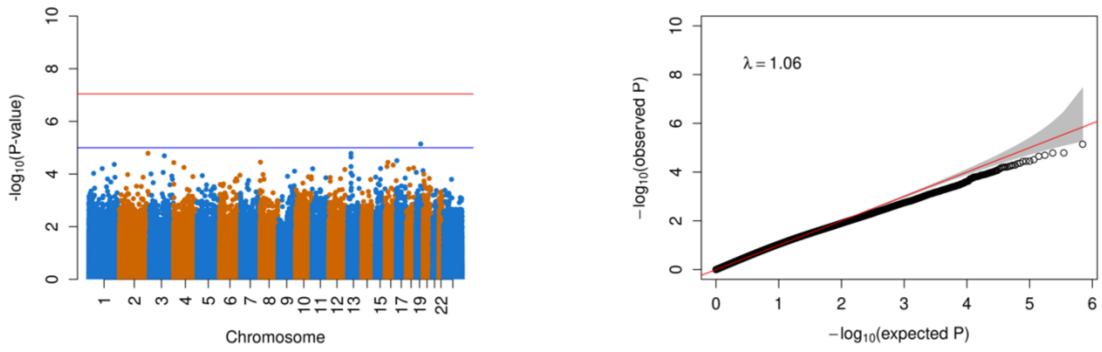

**B. CTH-adjusted EWAS**

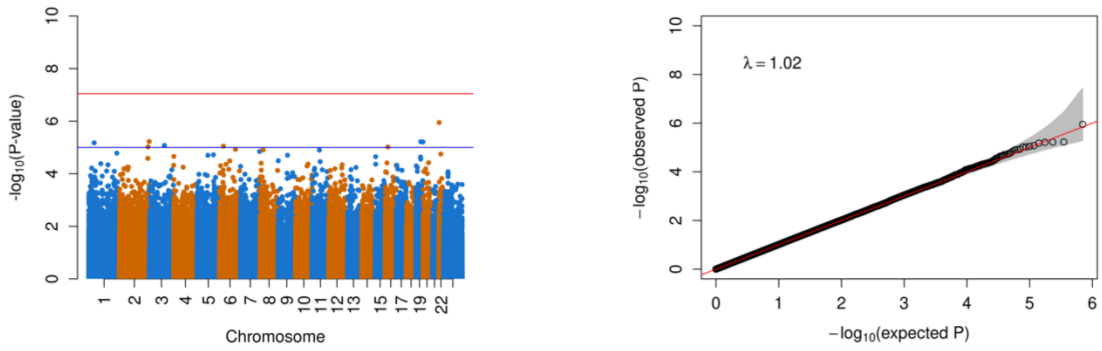

**C. CTH-unadjusted EWAS: sensitivity analysis**

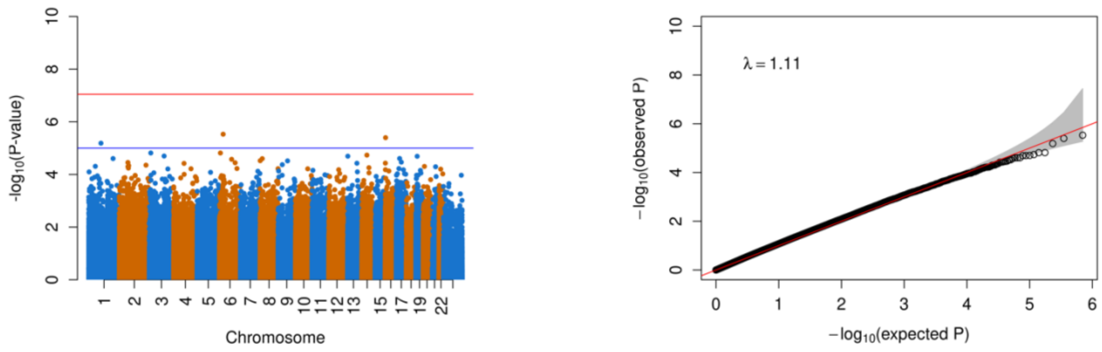

**D. CTH-adjusted EWAS: sensitivity analysis**

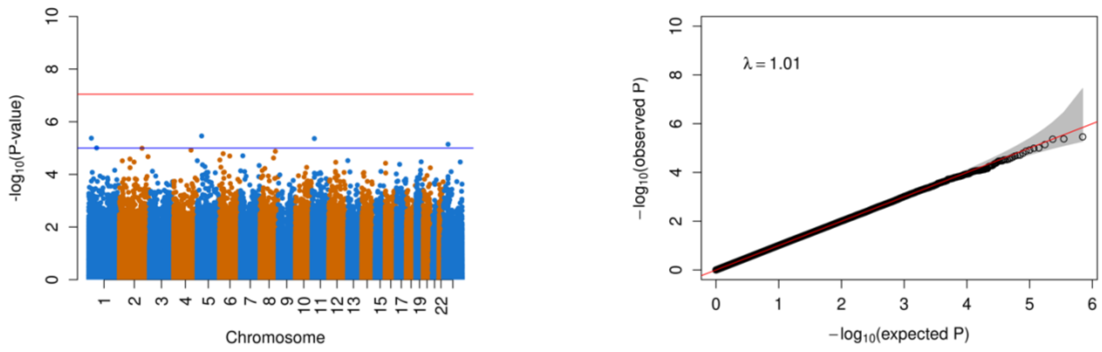

**Figure S11f. EWAS results for working memory**

**A. CTH-unadjusted EWAS**

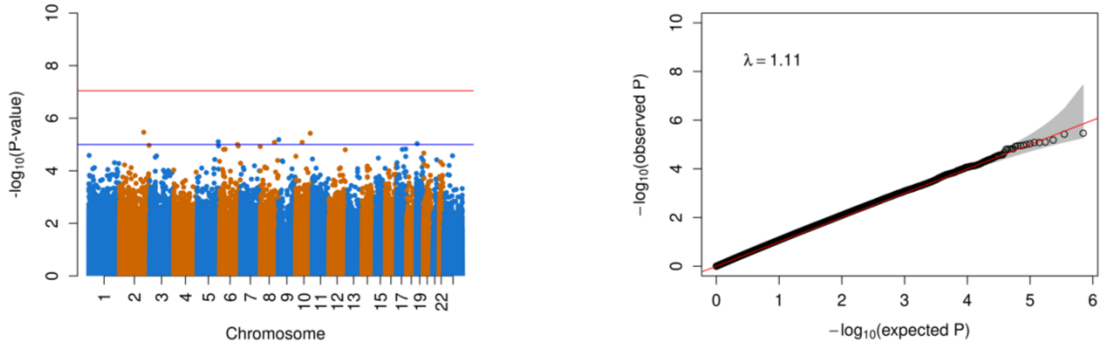

**B. CTH-adjusted EWAS**

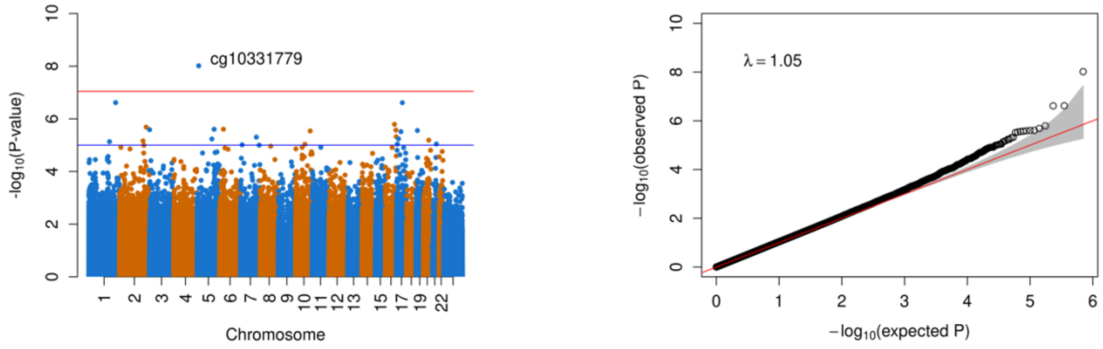

**C. CTH-unadjusted EWAS: sensitivity analysis**

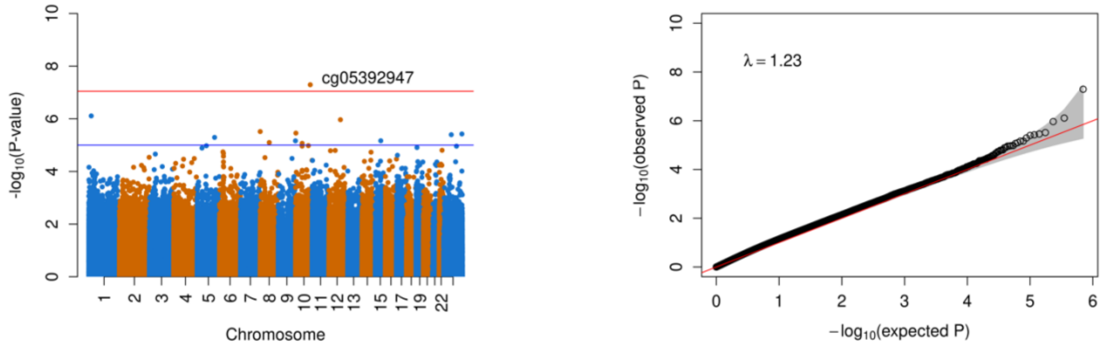

**D. CTH-adjusted EWAS: sensitivity analysis**

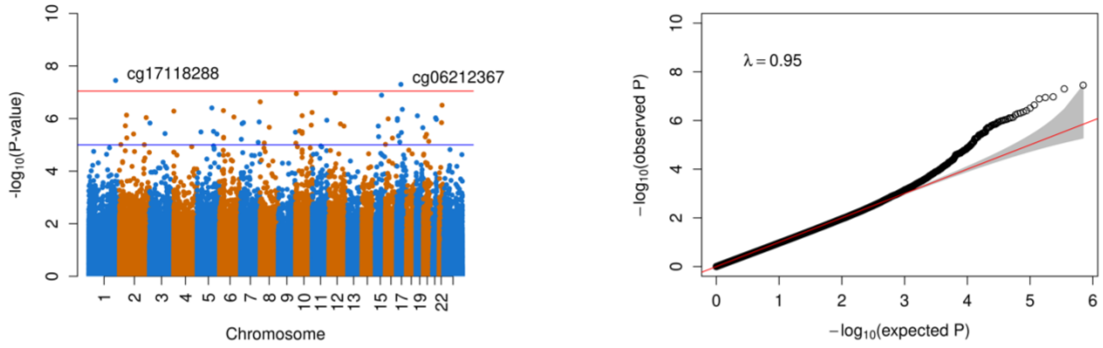

**Figure S11g. EWAS results for processing speed**

#### PAOFI total scores

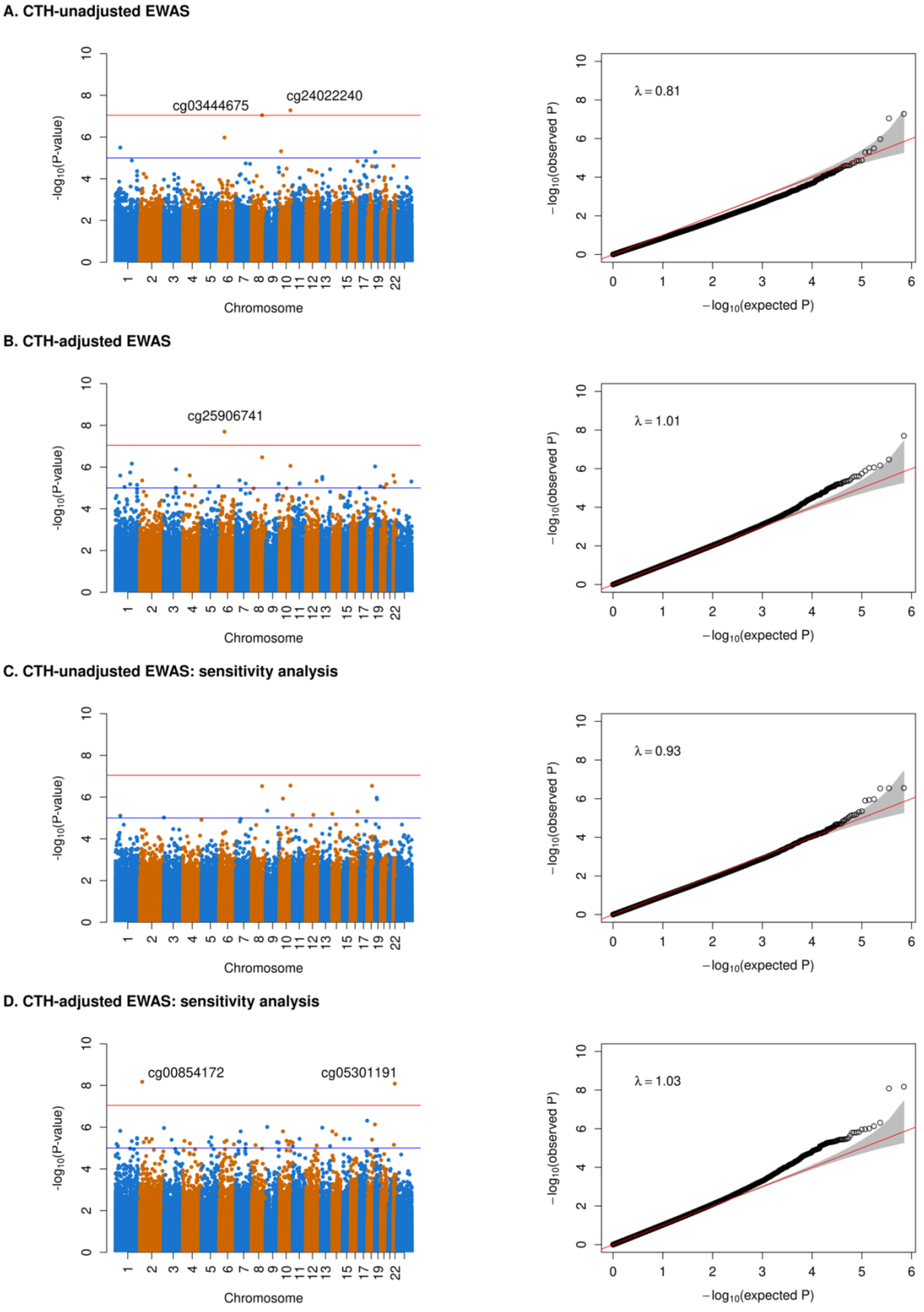

**Figure S11h. EWAS results for PAOFI total scores**

**Sensitivity analyses results**

Sensitivity analyses were conducted for a subset of samples who started the first ET treatment after the collection of blood samples. In the CTH-unadjusted EWAS of processing speed, cg05392947 in chromosome 10 with the nearest gene of *EMX2* was significant with p-value of $5.09 \times 10^{-8}$. cg17118288 in gene *TGFB2* on chromosome 1 (p-value = $3.55 \times 10^{-8}$) and cg06212367 in gene *SYNRG* on chromosome 17 (p-value = $5.01 \times 10^{-8}$) were significantly associated with processing speed with adjustment of surrogate variables controlling CTH. In the CTH-adjusted EWAS of PAOFI total scores, cg00854172 in gene *KLHL29* on chromosome 2 (p-value = $6.69 \times 10^{-9}$) and cg05301191 in gene *EIF4ENIF1* on chromosome 22 (p-value = $8.23 \times 10^{-9}$) were identified as significant DMCs. One significant DMR in *LIFR-AS1/LIFR* for processing speed was identified using summary statistics from CTH-unadjusted EWAS. With the adjustment of CTH, four DMRs for processing speed in/near gene *PRKG1/CSTF2T* (2), *TFCP2*, and *DOK4*, two DMRs for learning and memory in /near gene *CXXC4* and *SERINC4/C15orf63*, one DMR for verbal memory in *CCDC96*, and one DMR for PAOFI total scores in *SNHG22/MYO5B* were found. Next, GO and KEGG analyses were performed using results from CTH-adjusted EWASes. Two GO terms were identified for PAOFI total scores, including actin filament-based process and actin cytoskeleton organization.

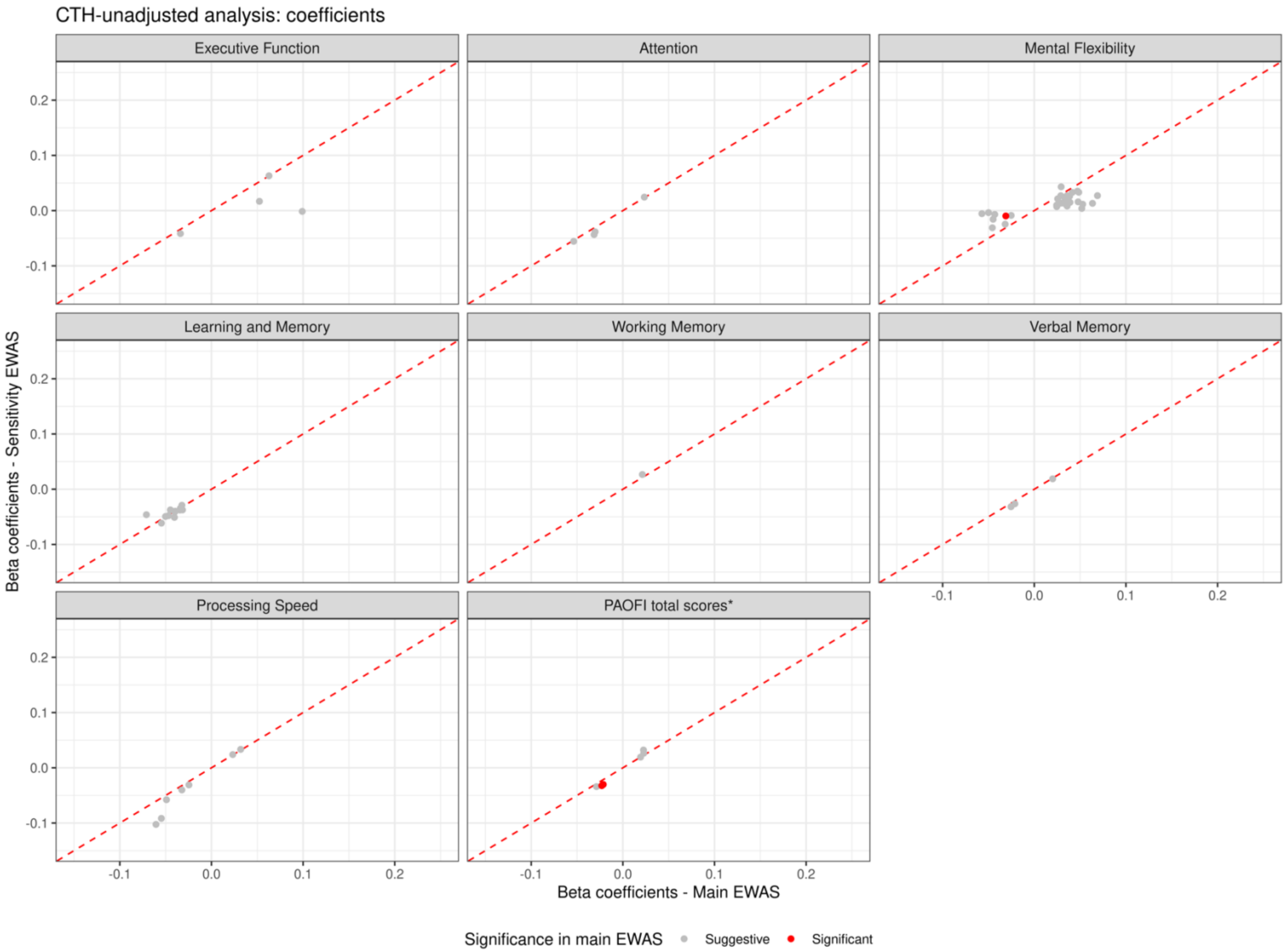

**Figure S12. Coefficients comparison between the main and sensitivity CTH-unadjusted EWASes**

Suggestive results from the main CTH-unadjusted EWAS (p-value $< 1 \times 10^{-5}$) were shown here. Suggestive CpGs were colored in grey and significant CpGs were shown in red.

*Since PAOFI total scores were on a different scale from the rest of the cognitive function phenotypes, the tenfold coefficients were presented.

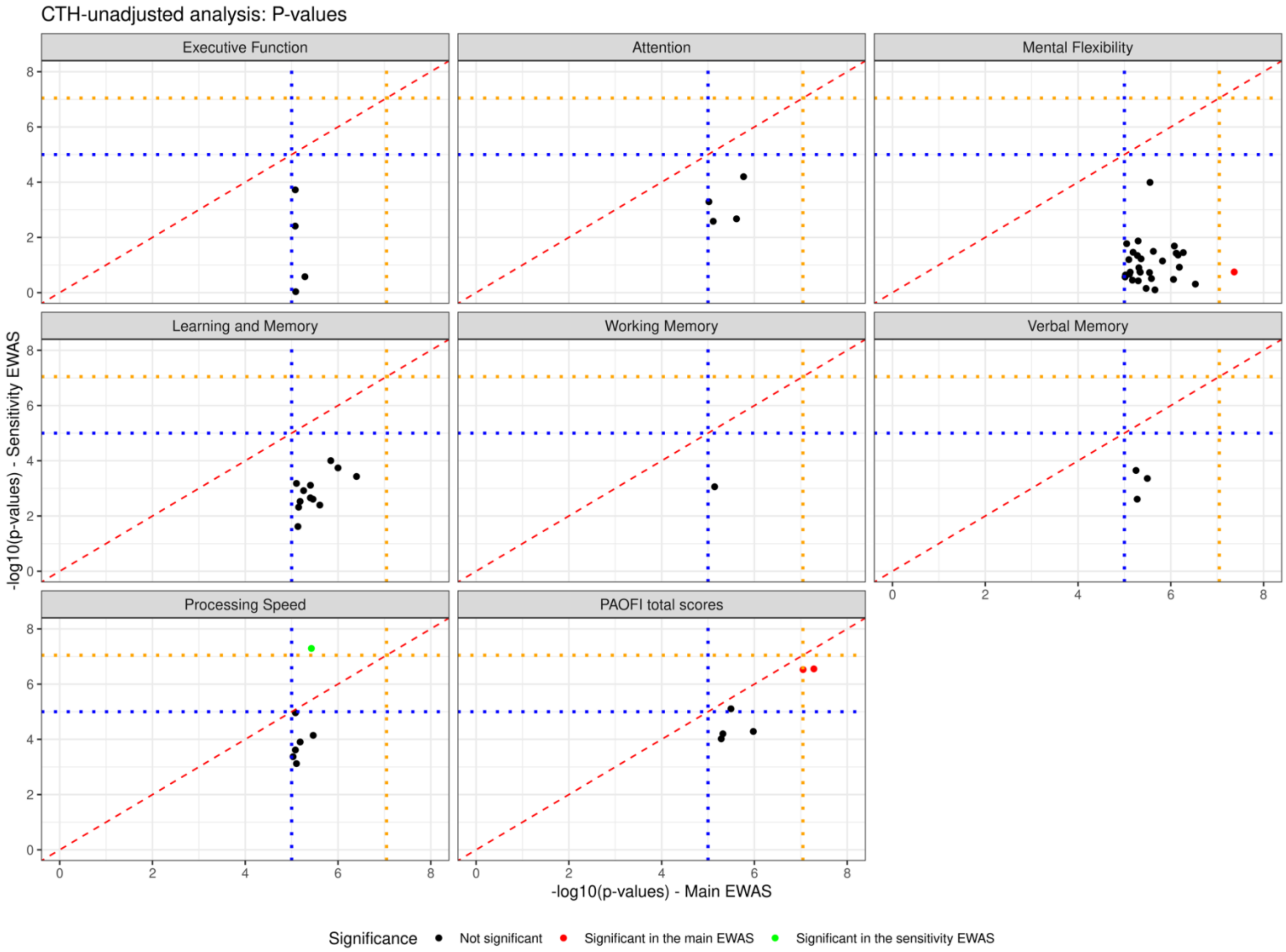

**Figure S13. P-values comparison between the main and sensitivity CTH-unadjusted EWASes**

Suggestive results from the main CTH-unadjusted EWAS (p-value < $1 \times 10^{-5}$) were shown here. The orange dashed line represents the genome-wide significance threshold of p-value = $9 \times 10^{-8}$ and the blue dashed line represents the suggestive threshold of p-value = $1 \times 10^{-5}$. Significant CpGs in the main EWAS were colored in red and significant CpGs in the sensitivity EWAS were colored in green.

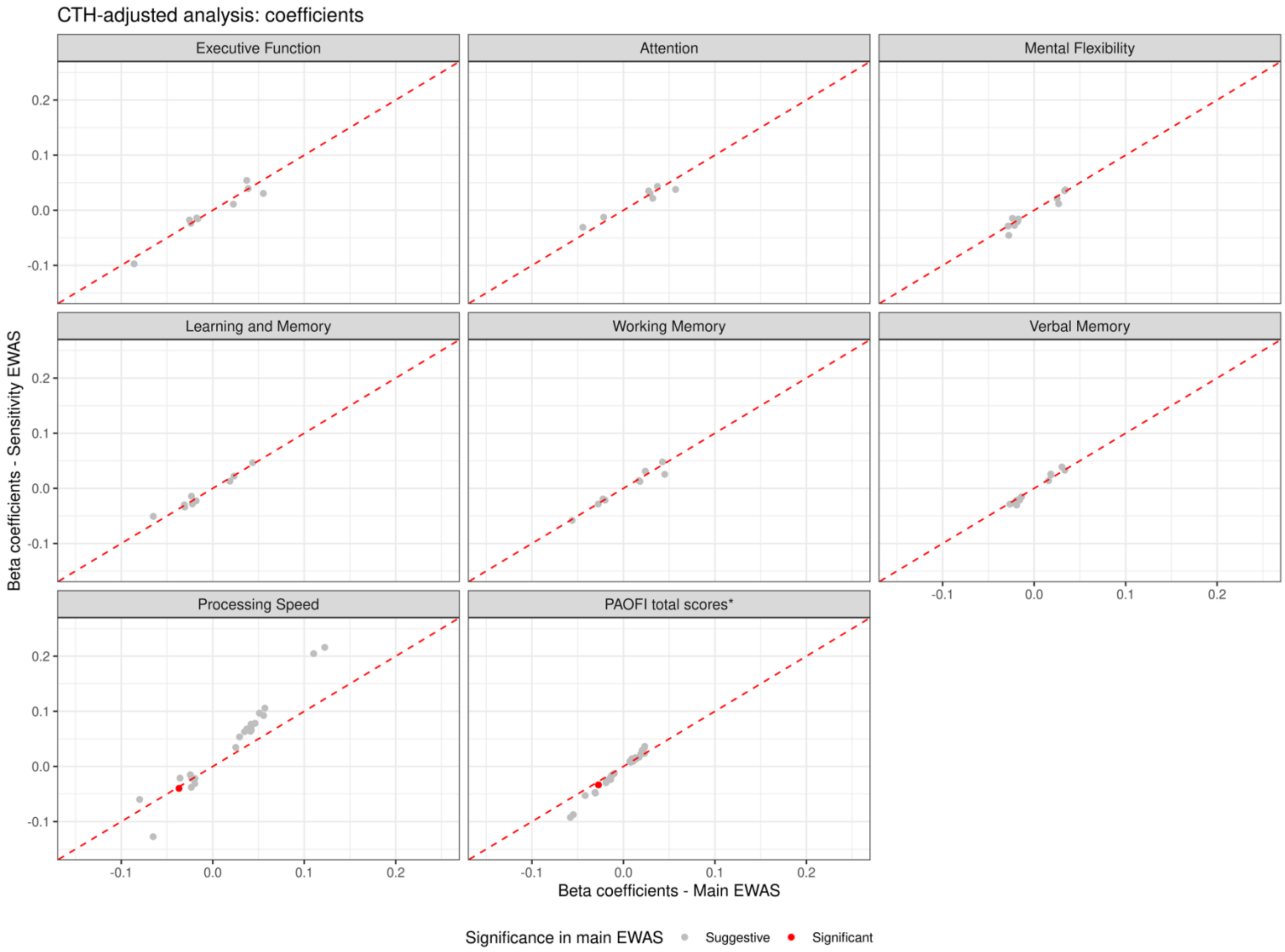

**Figure S14. Coefficients comparison between the main and sensitivity CTH-adjusted EWASes**

Suggestive results from the main CTH-adjusted EWAS (p-value $< 1 \times 10^{-5}$) were shown here. Suggestive CpGs were colored in grey and significant CpGs were shown in red.

*Since PAOFI total scores were on a different scale from the rest of the cognitive function phenotypes, the tenfold coefficients were presented.

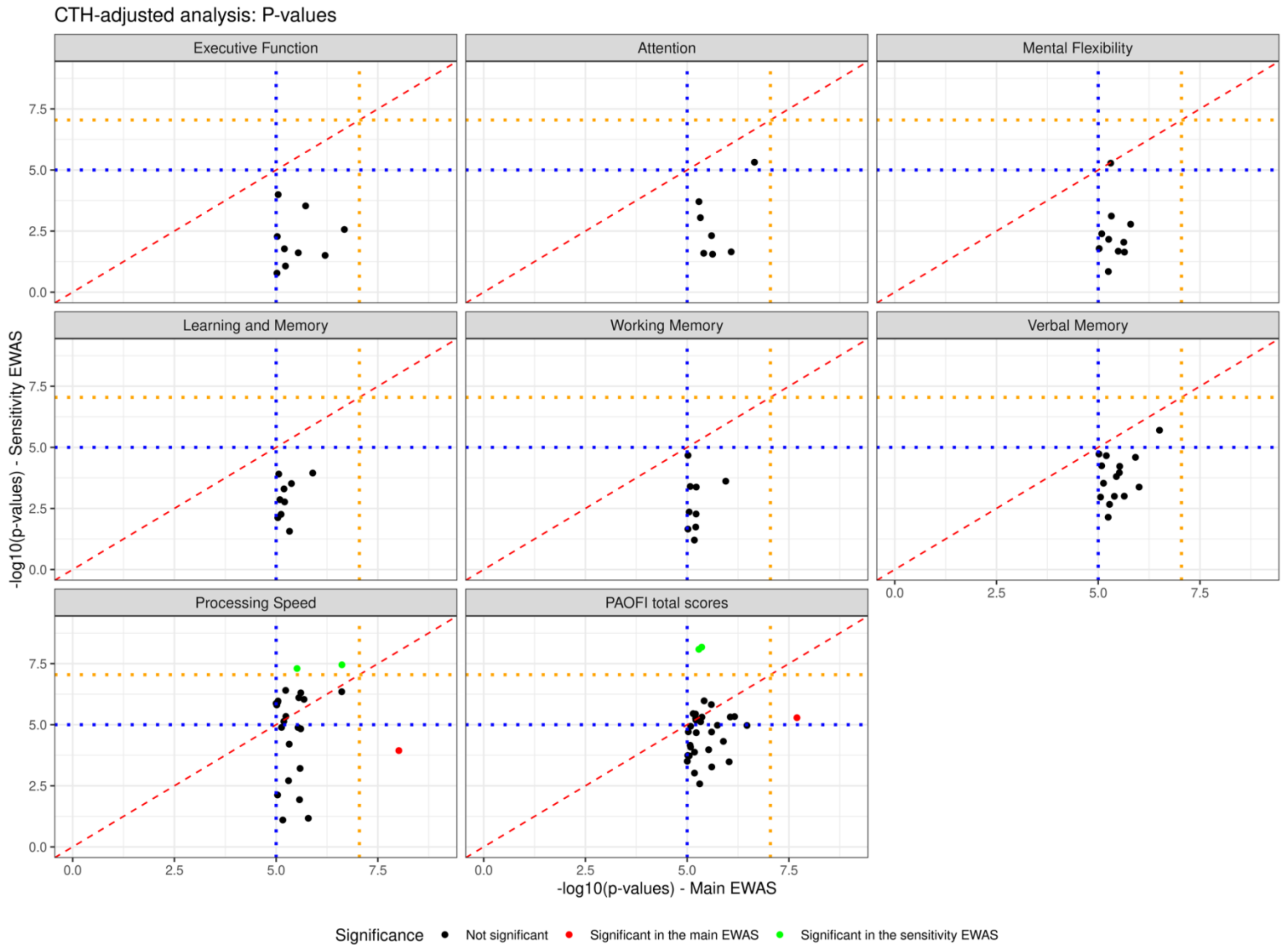

**Figure S15. P-values comparison between the main and sensitivity CTH-adjusted EWASes**

Suggestive results from the main CTH-adjusted EWAS (p-value $< 1 \times 10^{-5}$) were shown here. The orange dashed line represents the genome-wide significance threshold of p-value $= 9 \times 10^{-8}$ and the blue dashed line represents the suggestive threshold of p-value $= 1 \times 10^{-5}$. Significant CpGs in the main EWAS were colored in red and significant CpGs in the sensitivity EWAS were colored in green.

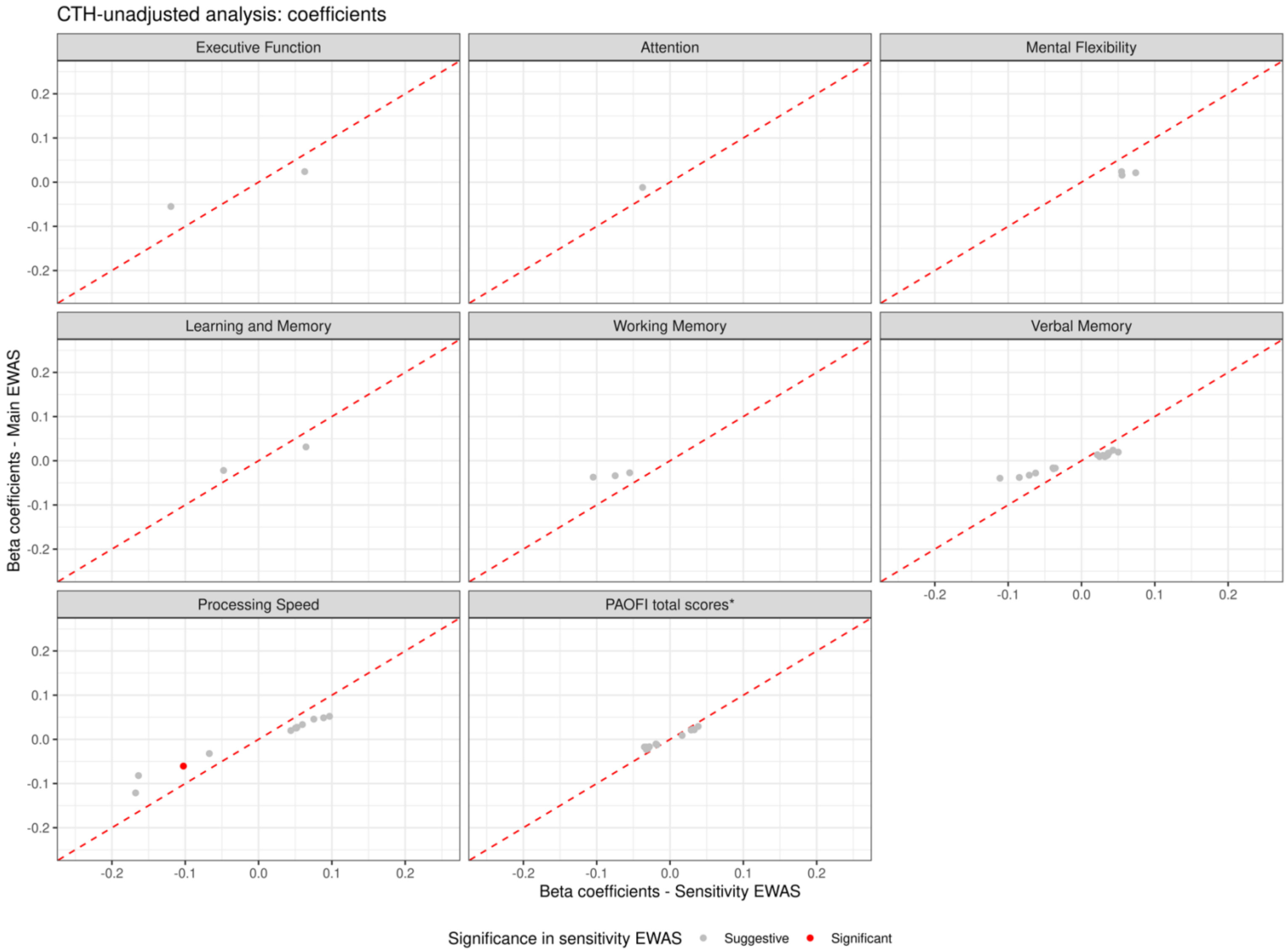

**Figure S16. Coefficients comparison between the main and sensitivity CTH-unadjusted EWASes**

Suggestive results from the sensitivity CTH-unadjusted EWAS (p-value $< 1 \times 10^{-5}$) were shown here. Suggestive CpGs were colored in grey and significant CpGs were shown in red.

*Since PAOFI total scores were on a different scale from the rest of the cognitive function phenotypes, the tenfold coefficients were presented.

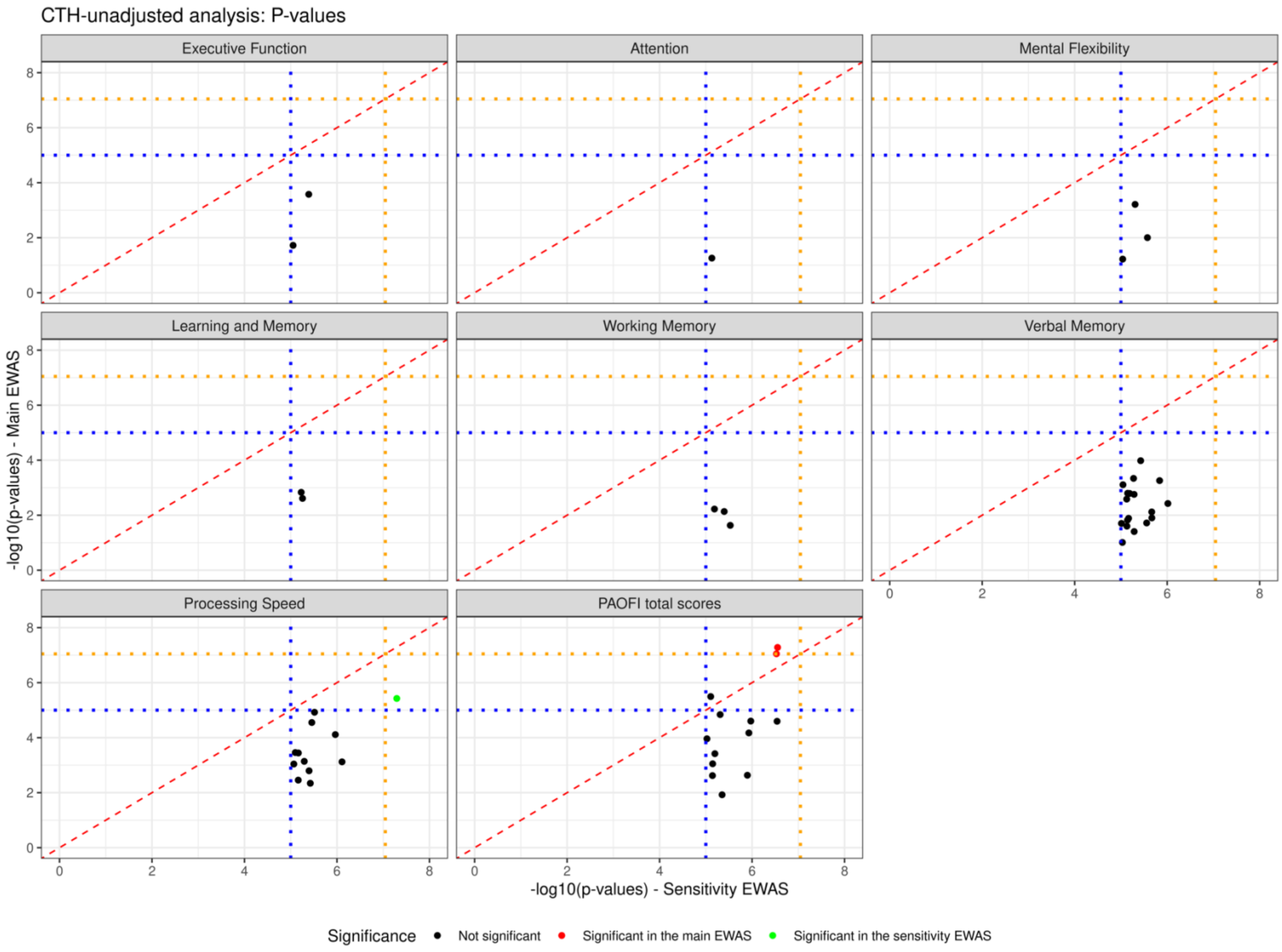

**Figure S17. P-values comparison between the sensitivity and main CTH-unadjusted EWASes**

Suggestive results from the suggestive CTH-unadjusted EWAS (p-value < $1 \times 10^{-5}$) were shown here. The orange dashed line represents the genome-wide significance threshold of p-value = $9 \times 10^{-8}$ and the blue dashed line represents the suggestive threshold of p-value = $1 \times 10^{-5}$. Significant CpGs in the main EWAS were colored in red and significant CpGs in the sensitivity EWAS were colored in green.

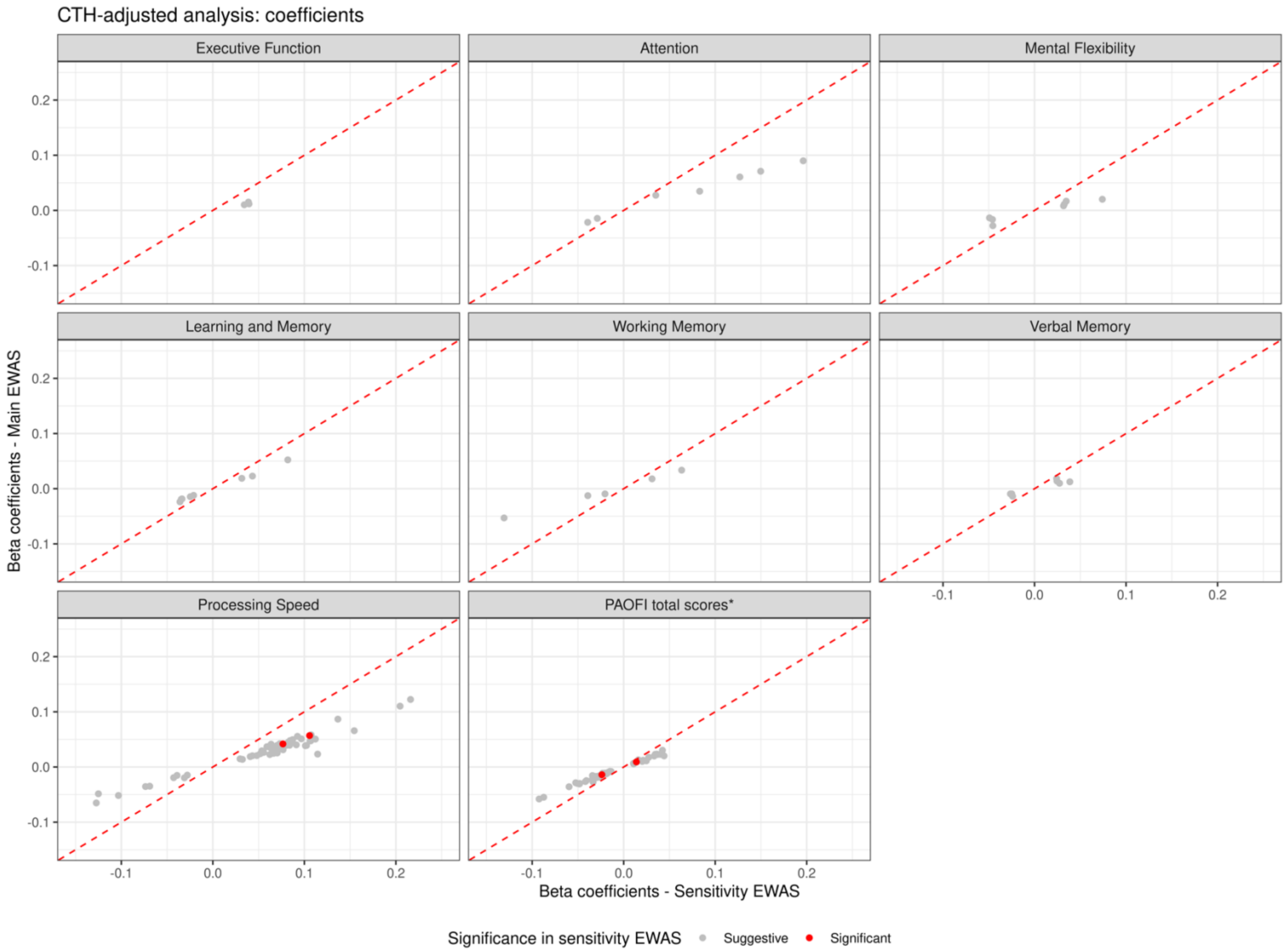

**Figure S18. Coefficients comparison between the sensitivity and main CTH-adjusted EWASes**

Suggestive results from the sensitivity CTH-adjusted EWAS (p-value $< 1 \times 10^{-5}$) were shown here. Suggestive CpGs were colored in grey and significant CpGs were shown in red.

*Since PAOFI total scores were on a different scale from the rest of the cognitive function phenotypes, the tenfold coefficients were presented.

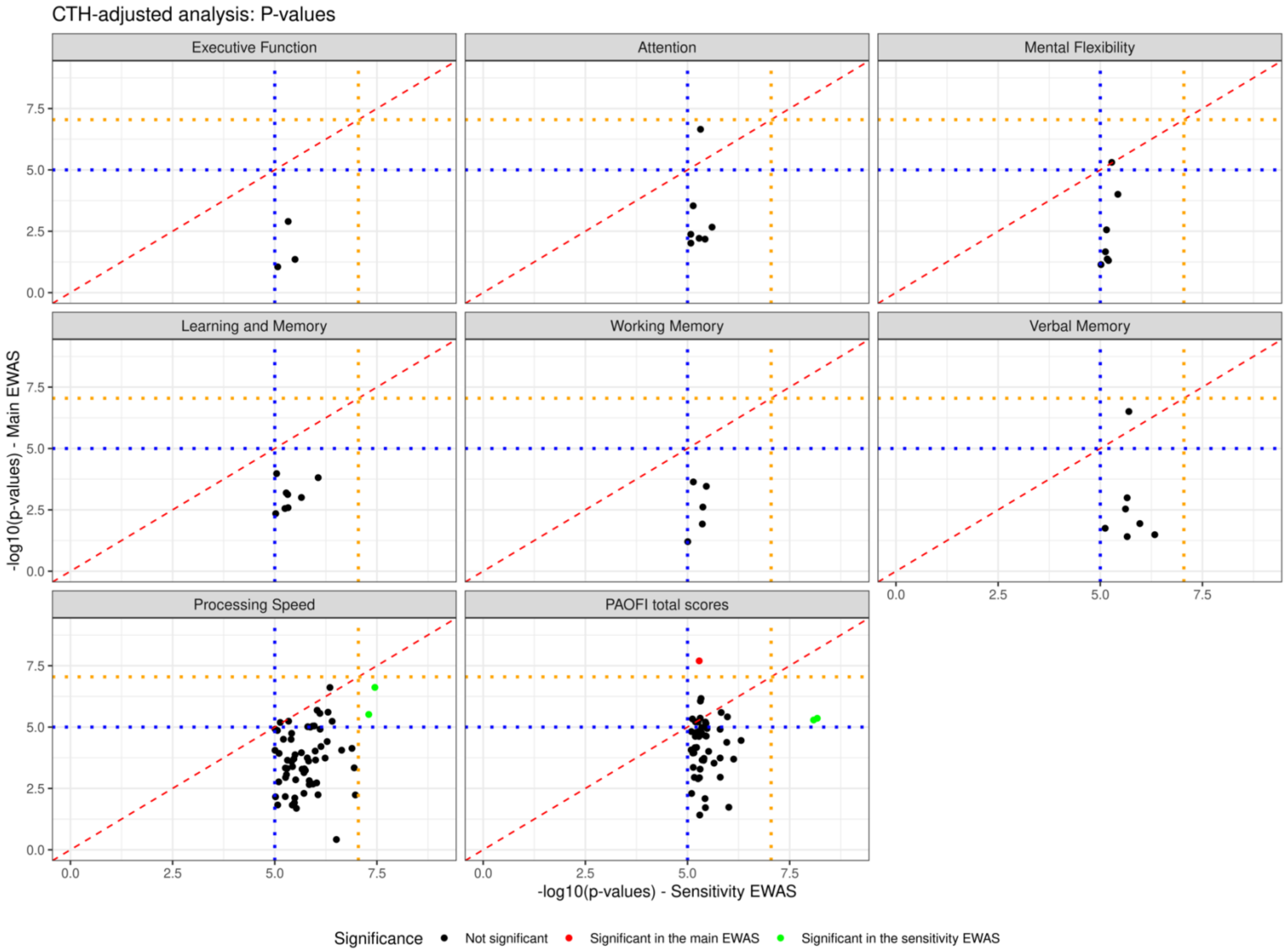

**Figure S19. P-values comparison between the sensitivity and main CTH-adjusted EWASes**

Suggestive results from the sensitivity CTH-adjusted EWAS (p-value < $1 \times 10^{-5}$) were shown here. The orange dashed line represents the genome-wide significance threshold of p-value = $9 \times 10^{-8}$ and the blue dashed line represents the suggestive threshold of p-value = $1 \times 10^{-5}$. Significant CpGs in the main EWAS were colored in red and significant CpGs in the sensitivity EWAS were colored in green.

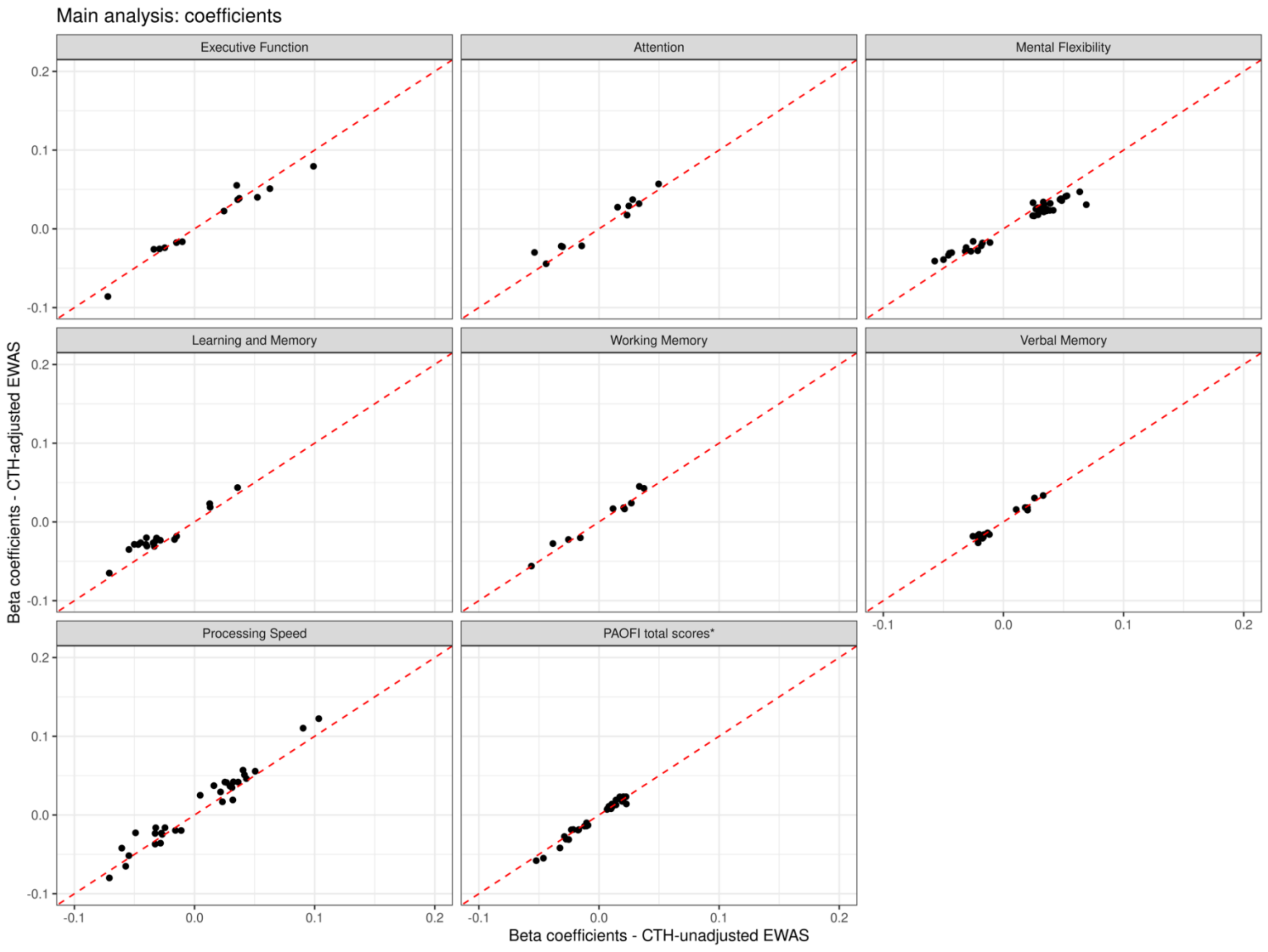

**Figure S20. Coefficients comparison between the main CTH-unadjusted and CTH-adjusted EWASes**

*Since PAOFI total scores were on a different scale from the rest of the cognitive function phenotypes, the tenfold coefficients were presented.

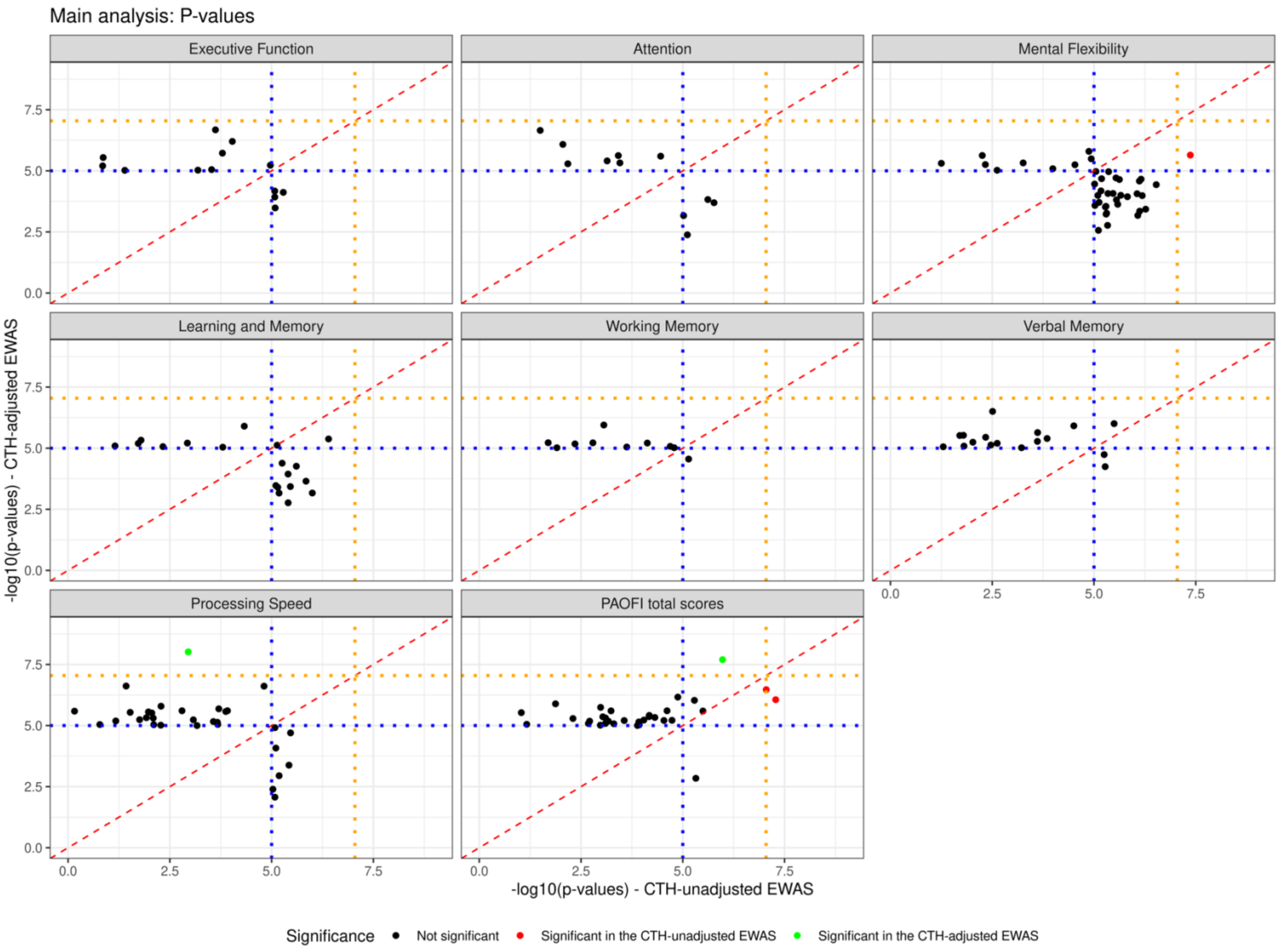

**Figure S21. P-values comparison between the main CTH-unadjusted and CTH-adjusted EWASes**

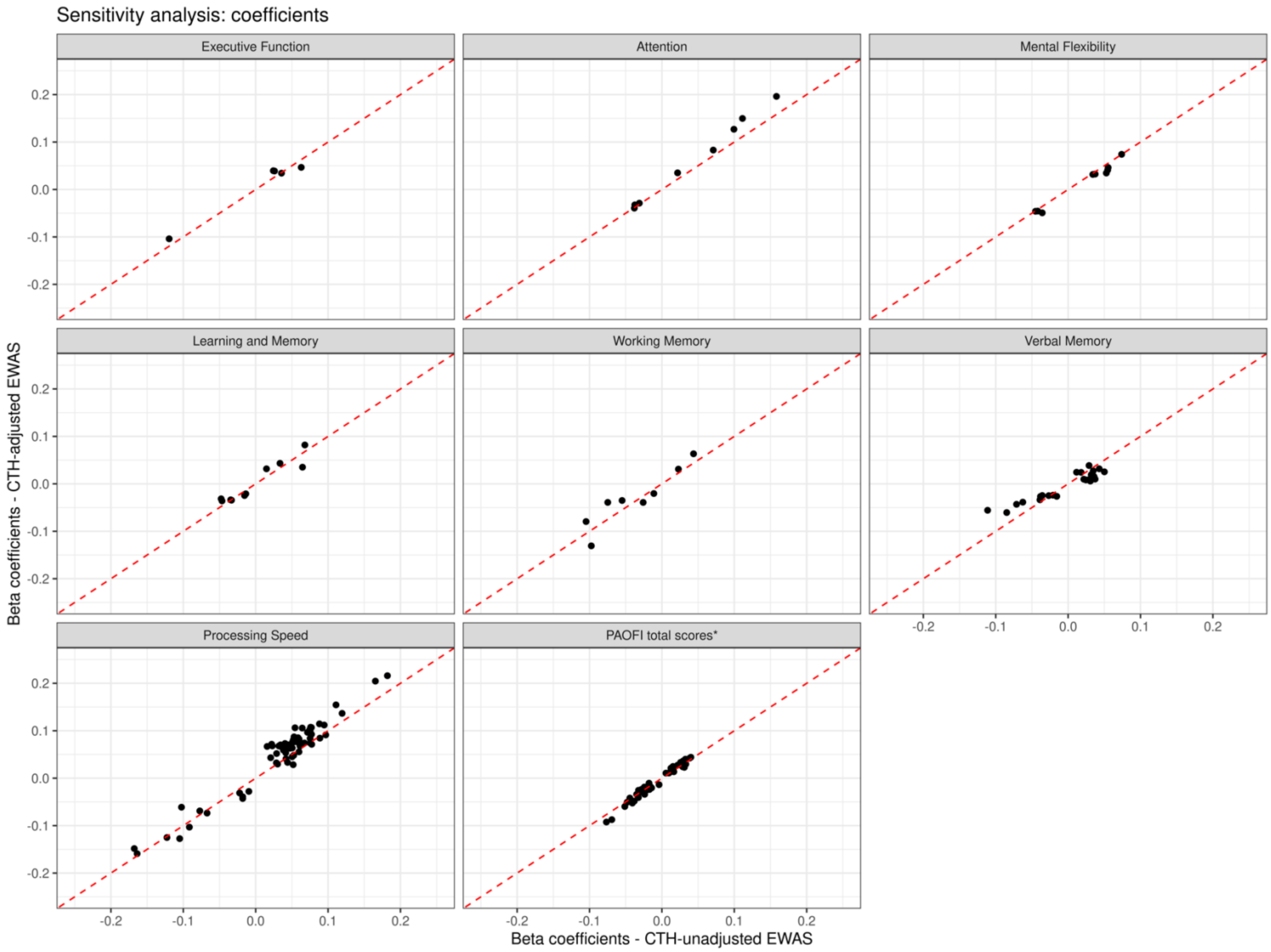

**Figure S22. Coefficients comparison between the sensitivity CTH-unadjusted and CTH-adjusted EWASes**

*Since PAOFI total scores were on a different scale from the rest of the cognitive function phenotypes, the tenfold coefficients were presented.

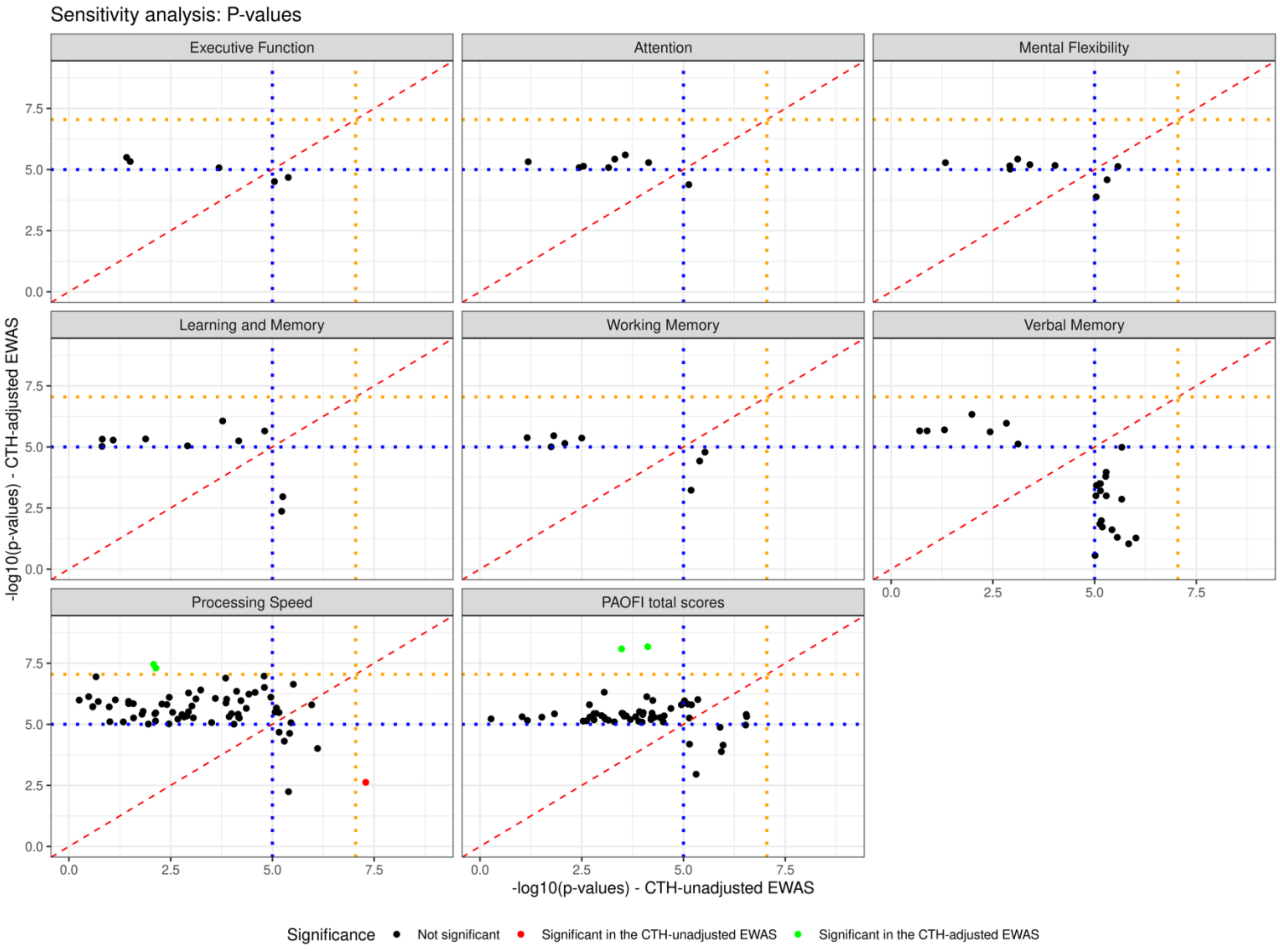

**Figure S23. P-values comparison between the sensitivity CTH-unadjusted and CTH-adjusted EWASes**

**A.**

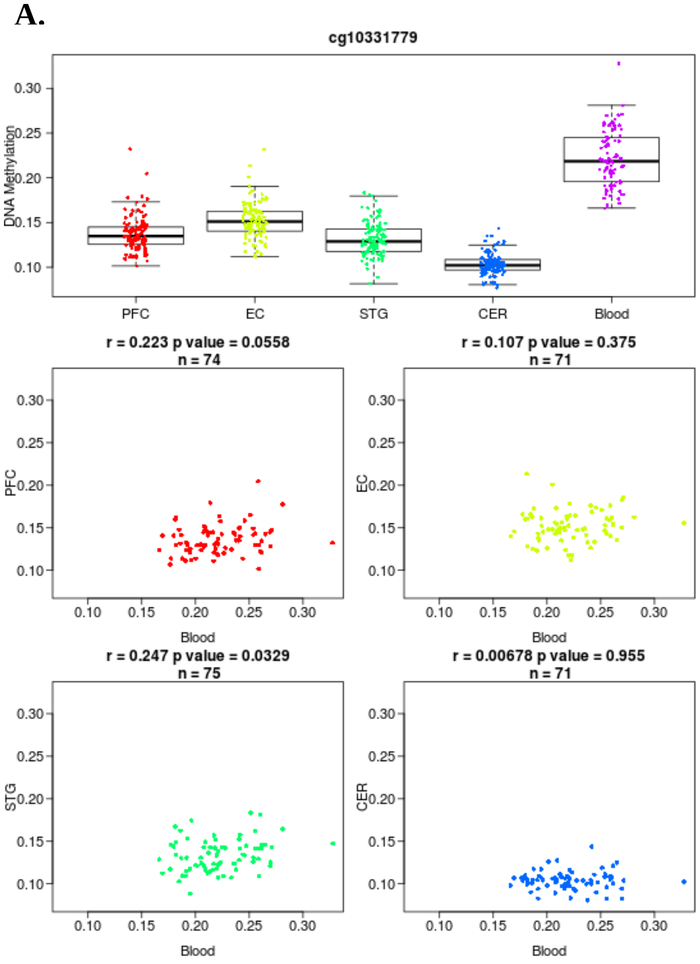

**B.**

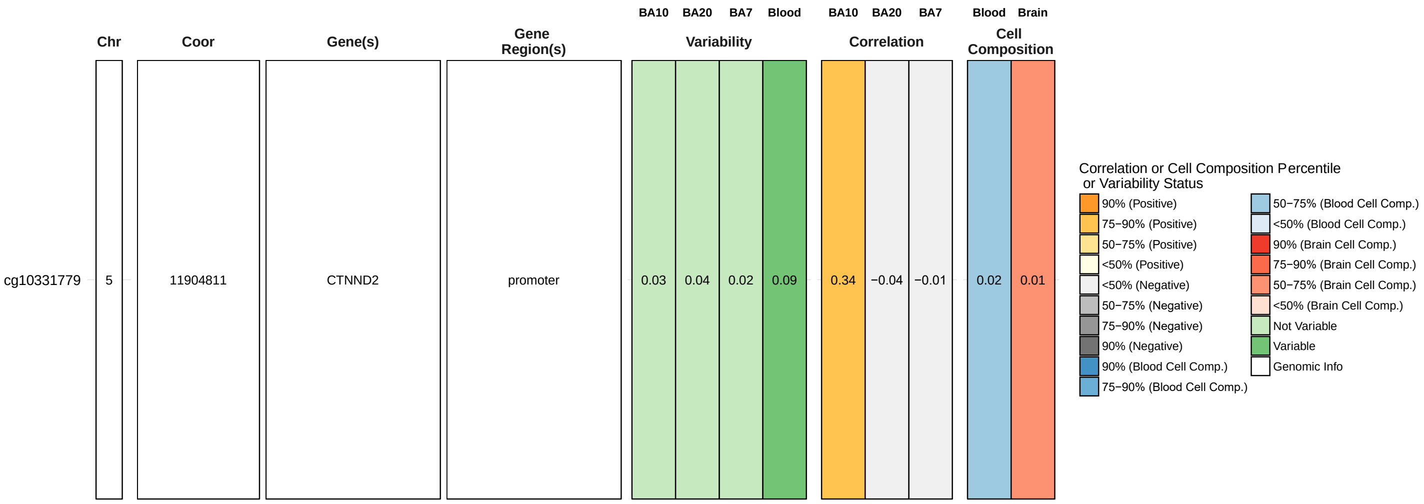

**Figure S24. cg10331779 DNAm correlation between blood and brain tissues**

A. Results from the blood brain DNA methylation comparison tool. PFC: prefrontal cortex, EC: entorhinal cortex, STG: superior temporal gyrus, CER: cerebellum.

B. Results from BECon. BA: Brodmann area.

**Table S3. Top DMCs of CTH-adjusted EWAS across all cognitive function phenotypes**

| CpG | Cognitive function phenotype | Effect size | P-value |
|---|---|---|---|
| **cg10331779** | | | |
| | Executive function | $-7.95 \times 10^{-3}$ | 0.247 |
| | Attention | $-8.32 \times 10^{-3}$ | 0.268 |
| | Mental flexibility | –0.014 | 0.018 |
| | Learning and memory | $-2.28 \times 10^{-3}$ | 0.710 |
| | Working memory | $6.08 \times 10^{-3}$ | 0.285 |
| | Verbal memory | $-7.64 \times 10^{-3}$ | 0.121 |
| | Processing speed | –0.037 | $\mathbf{9.65 \times 10^{-9}}$ |
| | PAOFI total scores | $3.94 \times 10^{-4}$ | 0.109 |
| **cg25906741** | | | |
| | Executive function | $1.27 \times 10^{-3}$ | 0.926 |
| | Attention | 0.015 | 0.331 |
| | Mental flexibility | $7.91 \times 10^{-3}$ | 0.515 |
| | Learning and memory | $-5.02 \times 10^{-3}$ | 0.681 |
| | Working memory | –0.020 | 0.086 |
| | Verbal memory | –0.011 | 0.284 |
| | Processing speed | $-7.93 \times 10^{-3}$ | 0.569 |
| | PAOFI total scores | $-2.73 \times 10^{-3}$ | $\mathbf{2.01 \times 10^{-8}}$ |

**Table S4. Lookup of top CTH-EWAS DMCs in McCartney *et al.* (2022)**

| CpG | Cognitive function phenotype* | Mean effect size | Standard error | PIP^ |
|---|---|---|---|---|
| **cg10331779** | | | | |
| | Digit symbol | $1.51 \times 10^{-5}$ | $1.70 \times 10^{-5}$ | 0.012 |
| | Logical memory | $-2.19 \times 10^{-6}$ | $2.77 \times 10^{-6}$ | 0.003 |
| | Verbal fluency | $8.73 \times 10^{-6}$ | $8.59 \times 10^{-6}$ | 0.006 |
| | Vocabulary | $3.24 \times 10^{-5}$ | $1.67 \times 10^{-5}$ | 0.007 |
| | General fluid cognitive function | $-1.32 \times 10^{-5}$ | $1.27 \times 10^{-5}$ | 0.011 |
| | General cognitive function | $-1.36 \times 10^{-5}$ | $2.27 \times 10^{-5}$ | 0.012 |
| **cg25906741** | | | | |
| | Digit symbol | $1.47 \times 10^{-5}$ | $1.40 \times 10^{-5}$ | 0.010 |
| | Logical memory | $-1.32 \times 10^{-5}$ | $7.17 \times 10^{-5}$ | 0.012 |
| | Verbal fluency | $1.37 \times 10^{-5}$ | $1.18 \times 10^{-5}$ | 0.007 |
| | Vocabulary | $-1.56 \times 10^{-5}$ | $2.25 \times 10^{-5}$ | 0.010 |
| | General fluid cognitive function | $-1.56 \times 10^{-5}$ | $1.70 \times 10^{-5}$ | 0.013 |
| | General cognitive function | $-2.13 \times 10^{-5}$ | $2.11 \times 10^{-5}$ | 0.013 |

* The general fluid cognitive function was derived from the first unrotated principal component of digit symbol, logical memory, and verbal fluency. The general cognitive function was derived from the first unrotated principal component of digit symbol, logical memory, verbal fluency, and vocabulary[5].
^ PIP: posterior inclusion probability

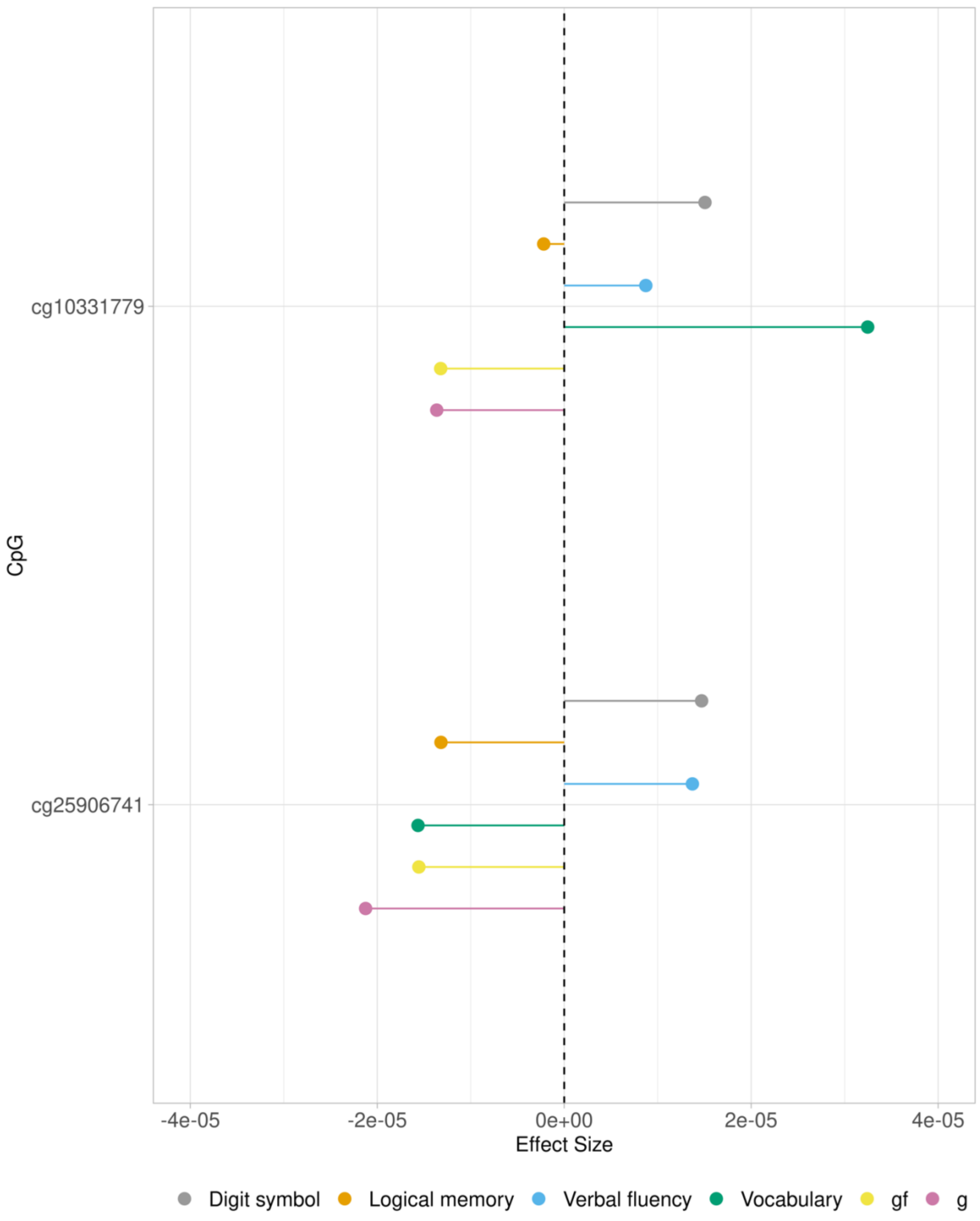

**Figure S25. Lookup EWAS associations in McCartney *et al.* (2022) for top DMCs cg10331779 and cg25906741**

gf: general fluid cognitive function, g: general cognitive function
